# Prenatal Alcohol Exposure and Mental Health Outcomes: A Two-Sample Mendelian Randomization Study of DNA Methylation Signatures

**DOI:** 10.64898/2026.01.30.26345158

**Authors:** Dailin Luo, Alexandre A. Lussier

## Abstract

Prenatal alcohol exposure (PAE) can lead to a range of deficits falling under the umbrella of Fetal Alcohol Spectrum Disorder (FASD), which included higher risk for adverse neurodevelopmental and mental health outcomes. Although the biological mechanisms underlying the link between PAE and mental health remain unclear, DNA methylation (DNAm), an epigenetic modification responsive to environmental exposures, may explain these relationships. Here, we applied a two-sample Mendelian randomization (MR) framework to assess whether DNAm loci previously associated with PAE or FASD are linked to 11 psychiatric outcomes. Using summary statistics from the Genetics of DNA Methylation Consortium (GoDMC) mQTL database and large-scale GWAS, we analyzed DNAm loci from two epigenome-wide association studies: one examining FASD by Lussier et al. (2018) and one examining PAE patterns by Sharp et al. (2018). A total of 106 associations (Lussier) and 28 associations (Sharp) reached nominal significance (p<0.05) and passed sensitivity tests, with several surviving multiple testing correction. Notably, schizophrenia and bipolar disorder had the highest number of associated loci across both studies. Functional analysis showed that DNAm loci were enriched in signaling pathways, embryonic development, and neuron differentiation. Regional enrichment analysis revealed that FASD-related loci were more likely to occur in enhancer and south shore, implicating distal regulatory elements. PAE patterns conferred heterogeneous effects on DNAm and mental health risk, underscoring the complexity of timing-specific epigenetic vulnerability. These findings offer novel insights into the potential mechanism of DNAm linking PAE to mental health, and demonstrate the utility of MR in epigenetic epidemiology.

## INTRODUCTION

Prenatal alcohol exposure (PAE) can disrupt brain development and cause deficits in cognitive, behavioral, and motor functions [1]. These deficits are known as fetal alcohol spectrum disorder (FASD), which includes fetal alcohol syndrome (FAS), partial fetal alcohol syndrome (pFAS), alcohol related neurodevelopmental disorders (ARND), and alcohol related birth defects (ARBD) [2]. Globally, FASD affects an estimated 7.7 cases per 1,000 individuals [3], with prevalence in the United States ranging from 1% to 5% among school-aged children [4–6]. Although often treated as a uniform exposure, the effects of PAE vary by timing and drinking pattern. For instance, Goldschmidt et al. [7] found that heavy first-trimester exposure was associated with offspring alcohol use and an 84% increase in two or more alcohol use disorder (AUD) symptoms after adjusting for other covariates, further highlighting the importance of examining pattern-specific relationships between PAE and mental health outcomes.

Beyond AUD, PAE is linked to a broad range of mental health disorders. Zhang *et al.,* reported that PAE significantly increased the risk of depression in offspring, (OR = 2.28, 95% CI: 1.61–3.25, *p <* 0.001) [8]. Other clinical and population studies have also consistently shown a high psychiatric burden across the lifespan for those with PAE and FASD. Among children, 87% met criteria for a psychiatric disorder [9], while nearly 90% of adults reported a diagnosis, most commonly ADHD (65%) and depression (47%) [10]. Within this prevalence, adolescents face particularly severe risks, with 35% reporting suicidal ideation and 13% at least one attempt in the past year [11]. Furthermore, high rates of substance misuse further reflect this vulnerability, with 38% misusing alcohol and 46% other drugs [12]. Collectively, these findings highlight the urgent need to clarify how PAE contributes to psychiatric vulnerability.

While the etiology of mental health in FASD remains unclear, recent evidence suggests alterations to DNA methylation (DNAm) have emerged as a potential pathway. DNAm regulates gene expression through the addition of methyl groups to cytosine of cytosine-guanine dinucleotides (CpG) sites, typically reducing transcription in promoter regions and enhancing it in gene bodies [13–14]. DNAm levels can also be shaped by genetic variants, such as single nucleotide polymorphisms (SNPs) that regulate specific CpG sites, referred to as methylation quantitative trait loci (mQTLs), and have been implicated in the architecture of complex traits [15]. Perhaps most importantly, DNAm is responsive to environmental factors, with multiple epigenome-wide association studies (EWAS) linking prenatal exposure to metals, tobacco smoke, and other factors to DNAm changes in newborns [16–18]. EWAS focused specifically on PAE or FASD and DNAm have reported mixed results [19]: Sharp et al. [20] found no significant associations between PAE and DNAm at false discovery rate (FDR) *< .*05., whereas, Portales-Casamar et al. [22] identified 658 FASD-associated CpG sites at FDR*< .*05, of which Lussier et al. [21] verified 161 in an independent cohort (FDR*< .*05). Moreover, differentially methylated CpG sites have been found in multiple psychiatric disorders, including schizophrenia, bipolar disorder, depression, etc. [23–25].

Based on these studies, we hypothesize that DNAm is a mechanism through which PAE influences mental health outcomes. To test this hypothesis, we investigated existing PAE-DNAm associations and their relationship with psychiatric outcomes using two-sample Mendelian Randomization (MR). MR has become a popular approach for causal inference and has been widely applied in epidemiological studies (reviewed in [26]). It leverages genetic variants, randomly assigned at conception according to Mendel’s law of inheritance, and thus being independent of environmental confounders [27]. This method is especially valuable when randomized controlled trial (RCT) are not feasible or ethical, and when observational studies face challenges of unmeasured confounding bias and reverse causation [28]. Specifically, MR has previously been applied to examine the role of DNAm in relation to exposures such as tobacco smoke, prenatal smoke, vitamin B12, and glycemia (reviewed in [29]). To date, however, no studies have used two-sample MR analysis to investigate association between PAE-related DNAm and adverse mental health outcomes.

Overall, this study aimed to identify DNAm changes as potential mechanisms linking PAE to mental health disorders. To this end, we analyzed DNAm loci from two previous EWAS studies, Lussier et al., on FASD [21] and Sharp et al., on PAE [20], to investigate their associations with eleven distinct psychiatric outcomes, selected based on prior associations with PAE and/or FASD. To capture pattern-specific effects, the analysis on DNAm loci from Sharp et al., was conducted for five PAE patterns: drinking before pregnancy, binge drinking, sustained drinking, drinking during trimester one, and drinking during trimesters two or three. Further, functional analysis provided insight into the genes and biological processes implicated, while regional analyses identified the genomic regions where the DNAm loci are concentrated.

## METHODS

### Study Design

This study used a two-sample Mendelian Randomization (MR) framework to study the relationship between exposure, PAE- or FASD-related DNAm changes, and mental disorders in offspring, using single nucleotide polymorphisms (SNPs) as instrumental variables (IVs) (**Figure 1**).

**Figure 1:**
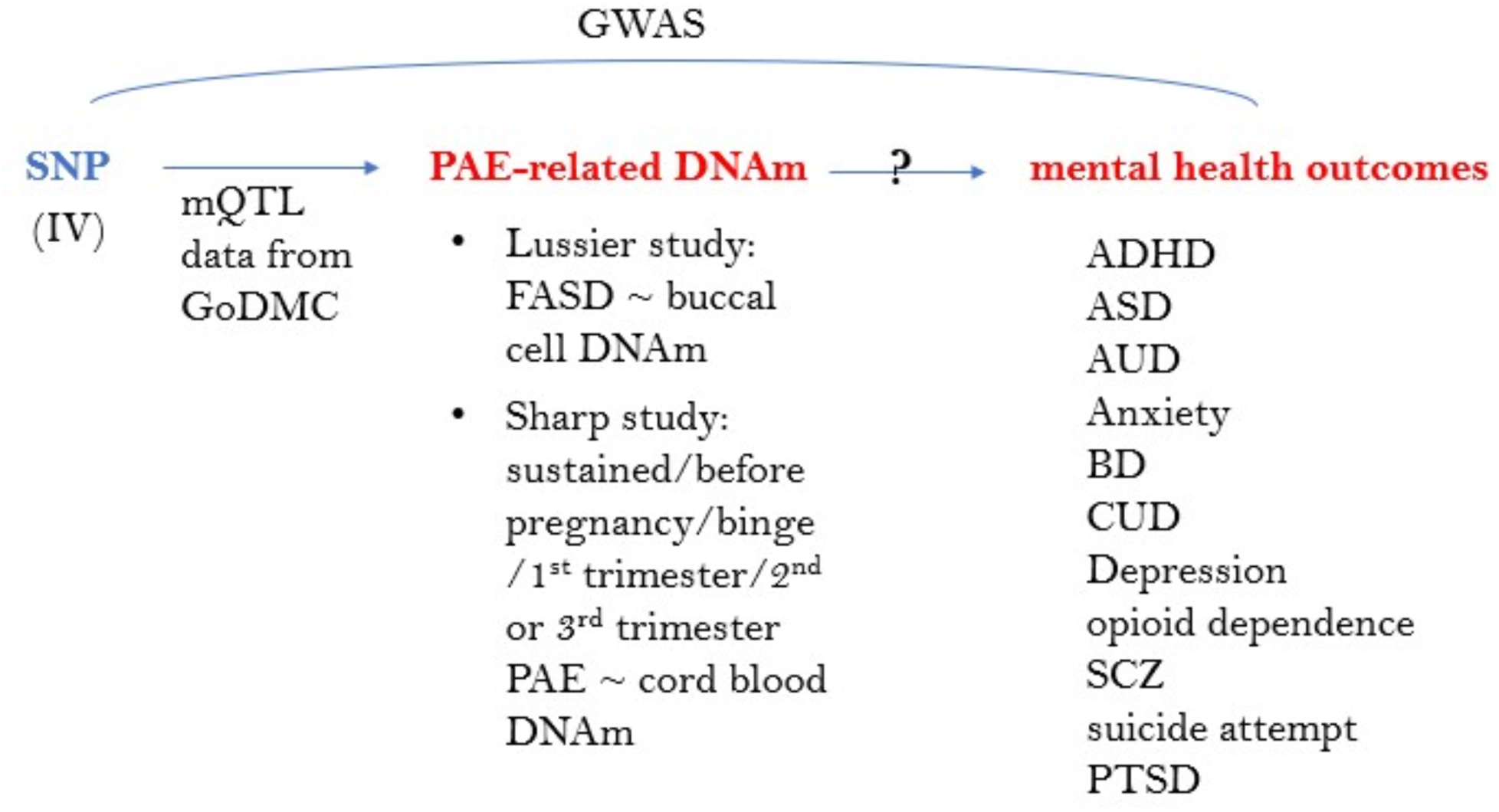
Schematic review of MR analysis. DNAm: DNA methylation; IV: instrumental variable; mQTL: methylation quantitative trait loci; ADHD: attention-deficit/hyperactivity disorder; ASD: autism spectrum disorder; AUD: alcohol use disorder; BD: bipolar disorder; CUD: cannabis use disorder; SCZ: schizophrenia; PTSD: post-traumatic stress disorder.

Although MR has many advantages, it relies on three assumptions:

- Relevance assumption: The SNPs being used as IVs must be robustly associated with the exposure.
- Independence assumption: The SNPs selected as IVs are not associated with confounders of the association between the exposure and the outcome.
- Exclusion restriction assumption (no horizontal pleiotropy assumption): There is no independent pathway between the IV and outcome other than through the exposure.

These assumptions were validated as described below in the section Two-Sample MR. The whole analysis consisted of the following steps: (1) obtaining SNPs that are significantly predictive for DNAm level changes of each CpG with the mQTL data from the Genetics of DNA Methylation Consortium (GoDMC), (2) extracting the instrument SNPs from the various genome-wide association studies (GWAS) of mental disorders, (3) harmonizing the effect sizes for the instrument SNPs on the exposure CpGs and the outcome to be each for the same reference allele, (4) performing MR analysis, (5) adjusting MR estimates of DNAm-outcome association for the direction of the PAE/FASD-DNAm association, (6) adjusting the p-values for multiple testing correction per outcome, and obtaining associations with p-values below a nominal threshold, (7) conducting sensitivity tests to filter out results that may have violated the assumptions of MR, (8) performing functional and regional analyses of nominal loci.

### EWAS of PAE- and FASD-related DNAm

DNAm changes associated with PAE or FASD were obtained from two epigenome-wide association studies (EWAS) (**Table S2**), both using the Illumina Human Methylation 450K array. Portales-Casamar et al. [22] initially analyzed DNA methylation data from buccal epithelial cells (BECs) collected from 110 FASD samples and 96 matched controls aged 5 to 18 years in the NeuroDevNet cohort. To account for ethnicity confounding, they conducted a second analysis on a more genetically homogeneous subgroup from the same cohort, consisting of 49 FASD samples and 87 controls, and eventually identified 658 differentially methylated CpG sites (*FDR < .*05). The subsequent study by Lussier et al. [21] validated these findings in 24 FASD and 24 control samples aged 3.5 to 18 years from the Kid Brain Health Network cohort, replicating altered methylation levels at 161 CpG sites (*FDR <* 0.05). The DNAm-FASD associations had been adjusted for batch effect, clinical status, and surrogate variables that may associate with known covariates, such as age and cell-type proportion. The cohorts of both studies were both Canadian-based. This manuscript analyzed the CpG sites identified in the Lussier study. As PAE leads directly to FASD, we assumed that FASD-associated DNAm changes can be used to gauge the effect of PAE on mental health outcomes through DNAm alterations.

The second study, Sharp et al. [20], evaluated impacts of different patterns of PAE (sustained alcohol consumption, binge drinking, drinking before pregnancy, drinking in the first trimester, drinking in the second or third trimester) on cord blood DNAm measured at birth. Although the authors reported no associations after multiple testing correction, 53 CpGs were associated with PAE with a less stringent threshold, p-value *<* 1×10^−5^. Among these, 8 CpGs were identified in sustained drinking, 25 in binge drinking, 5 in drinking before pregnancy, 3 in drinking during the first trimester, and 12 in drinking during the second or third trimester. The PAE-DNAm associations had been adjusted for covariates including maternal age, education, smoking status, and technical variables, either batch ID or surrogate variables. The cohorts that the Sharp study used (ALSPAC, GECKO, Generation R, MoBa1, MoBa2) were all based in Europe, consisting of 3075 mother-child pairs (1,147 PAE and 1,928 control samples).

### GWAS of DNAm

SNPs associated with the PAE- or FASD-related DNAm loci were retrieved from GoDMC database [30]. The GoDMC database includes DNAm quantitative trait locus (mQTL) data from blood samples of over 30,000 European participants, for a total of 420,509 loci. DNA methylation (DNAm) level data used in this study from GoDMC were assessed using the 450K array as continuous values between 0 and 1, which represents the fraction of cells with DNAm at a given locus.

### GWAS of Mental Health Outcomes

I focused on mental health outcomes previously identified in people with PAE or FASD.

In total, 11 health outcomes were selected, namely attention-deficit/hyperactivity disorder (ADHD), anxiety disorder, Autism spectrum disorder (ASD), alcohol use disorder (AUD), bipolar disorder (BD), cannabis use disorder (CUD), depression, opioid dependence, schizophrenia (SCZ), suicide attempts, and post-traumatic stress disorder (PTSD). The GWAS of AUD performed 3 analyses, for total score (AUD-T) of the Alcohol Use Disorders Identification Test (AUDIT), score of items 1-3 in AUDIT for alcohol consumption (AUD-C), and score of items 4-10 in AUDIT for alcohol-related problems (AUD-P). The GWAS for Opioid dependence compared opioid-dependent population to opioid-exposed population. Genetic data on summary level for all mental health outcomes were sourced from the largest publicly available GWAS (**Table 1**), all derived from European samples. Sample sizes varied from 3,272 cases and 2,876 controls for opioid dependence to 371,184 cases and 978,703 controls for depression. Study-specific covariates were adjusted for in each GWAS, such as sex, year of birth, genetic principal components, etc. All outcomes were coded as binary variables (ever or never), except for AUD-T, AUD-C, and AUD-P, which were coded continuously.

**Table 1:**
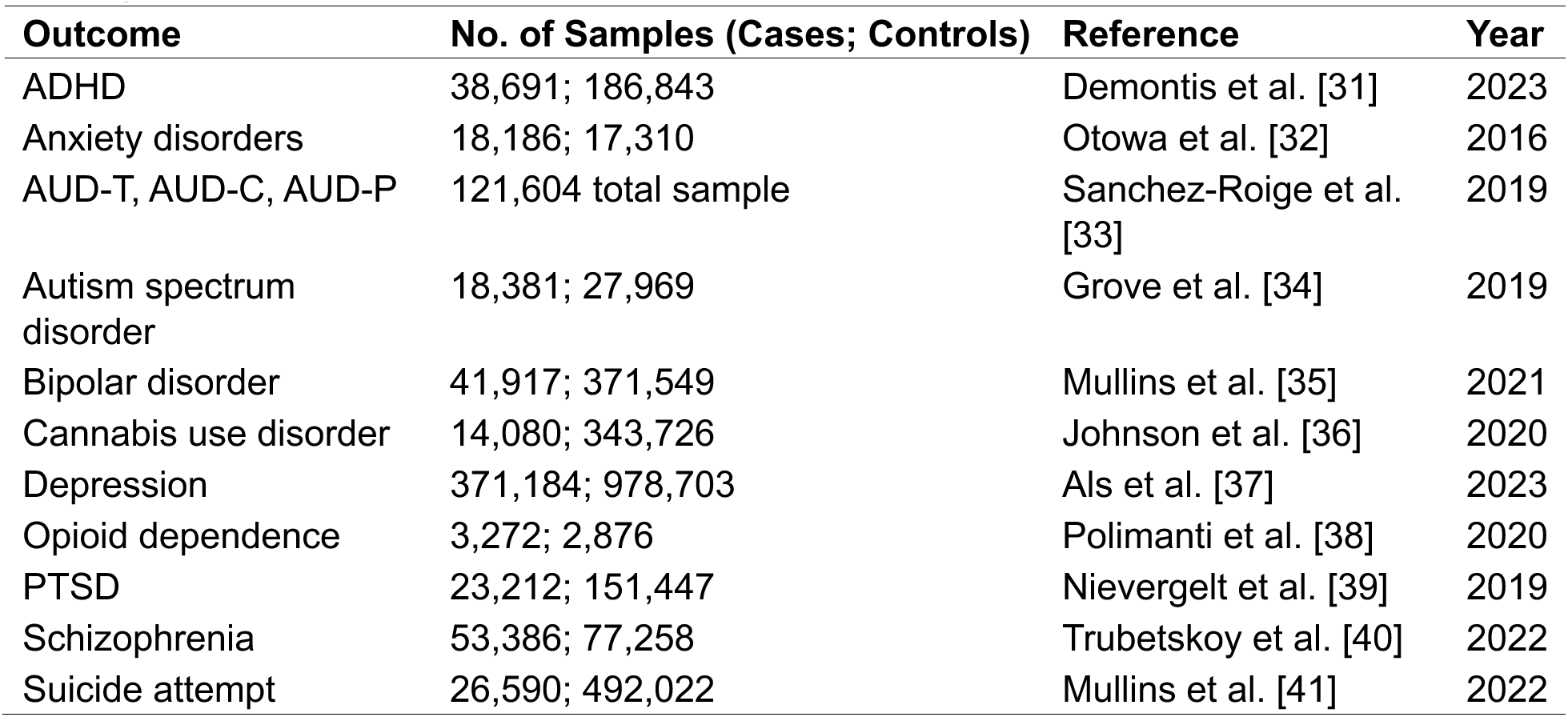
GWAS used to derive summary-level SNPs-outcome association data for each outcome. ADHD: attention-deficit/hyperactivity disorder; AUD-T/AUD-C/AUD-P: total score/consumption score/problem score of the Alcohol Use Disorders Identification Test (AUDIT); PTSD: post-traumatic stress disorder.

### Instrumental Variable Selection

We analyzed DNAm changes at loci previously associated with PAE or FASD (**Table S2**). In the Lussier study, all 161 DNAm loci were included, as they all had adjusted p-value below 0.05. In the Sharp study, 53 DNAm loci with p-value *<* 0.05 were used. After filtering, we identified SNPs associated with these DNAm loci either in cis (within 1 Mb from locus; p *<* 1 × 10^−8^) or trans (more than 1 Mb from locus; p *<* 1 × 10^−14^) position from the clumped data of GoDMC and extracted the SNP-DNAm associations. For these mapped SNPs, we retrieved their associations with the mental health outcomes from GWASs listed in **Table 1**. Associations between IVs and DNAm are presented as standardized effect estimates (z scores), reflecting the difference in DNAm level for each additional SNP allele. All IVs are listed in **Tables S3-S4**.

### Two-Sample Mendelian Randomization

We performed two-sample MR to study associations between DNAm and adverse mental health outcomes by using the TwoSampleMR package (version 0.6.11) in R (version 4.3.2). MR analyses were performed separately for each EWAS study, for each DNAm locus, and for each health outcome. To investigate the pattern-specific influences of PAE, MR was run separately for each pattern defined in the Sharp study. DNAm-outcome associations were estimated by Wald Ratio (WR) method, for DNAm loci with only 1 SNP available, or Inverse Variance Weighting (IVW) method, for those with more than 1 SNP available. If no SNP was available, the DNAm-outcome association analysis was skipped, due to a violation of the relevance assumption.

We reported associations with unadjusted p-values below 0.05 as nominal associations to facilitate discovery in downstream functional analyses that require larger numbers of loci. To address the potential issue of multiple testing, we also report p-values adjusted by Benjamini-Hochberg (BH) method that corrected for the number of loci tested in each health outcome, denoted as *FDR*.

To ensure the relevance assumption of MR, the p-values of the additive random effect (ARE) estimates between SNPs and DNAm loci were examined to ensure all the loci analyzed were robustly related to the SNPs to which they were mapped.

For the independence assumption, the mQTL data from GoDMC had been adjusted for sex, age, batch variables, smoking status, cell counts, genetic PCs, and nongenetic DNAm PCs, which limited the correlation between the SNPs and confounders that may also confound the DNAm-mental health outcome relationships. The two-sample design also helped ensure the independence assumption, by using one sample to retrieve summary data for SNP-exposure (i.e., DNAm) associations and another independent sample to retrieve summary data for SNP-outcome (i.e., mental disorder) associations.

For the exclusion restriction assumption, we used the expression quantitative trait methylation (eQTM) data on the HELIX Web Catalog (https://helixomics.isglobal.org/) to check if any DNAm loci were associated with gene expression in blood samples [42]. We also used the GWAS Catalog (https://www.ebi.ac.uk/gwas/) to determine whether any SNPs had known associations with the health outcomes in this study. Most importantly, we conducted three sensitivity tests after MR analysis to screen out the results that may violate the “no horizontal pleiotropy” assumption. These tests are:

- Heterogeneity test: we used Cochran’s Q statistic to assess whether different IVs provide consistent estimates. A *p < .*05 for the Q statistic was used as the criterion of failing the heterogeneity test. This test requires at least 2 SNPs to be performed.
- Pleiotropy test: we assessed whether IVs influence the outcome through pathways other than the exposure by checking whether the intercept of MR-Egger regression significantly deviated from zero. A *p < .*05 for the intercept was used as the criterion of failing the pleiotropy test. This test requires at least 3 SNPs to be performed.
- Leave-one-out test: we tested each DNAm locus identified as associated with a mental health outcome with more than one SNP (i.e., those had an overall *p < .*05 calculated by IVW method). If estimating by IVW while excluding one of the SNPs generated a *p > .*05, we considered this DNAm locus as having failed the leave-one-out test. In this way, we checked whether a single IV disproportionately affected the estimate.

This test requires at least 2 SNPs to be performed. Results that failed one or more sensitivity tests were excluded from downstream analysis.

### Functional Analysis

The missMethyl package (version 1.36.0) in R was used to map CpGs to the genes in which they are located, with the annotation data of the 450K array. The missMethyl package was also utilized to perform functional enrichment analysis for the risk-increasing and risk-suppressing DNAm loci, separately, after filtering out the MR results that failed one or more sensitivity test(s). The top 100 gene ontology (GO) terms were input into REVIGO (http://revigo.irb.hr/) [43]. REVIGO is a web tool that summarizes long lists of GO terms by removing redundant terms and finding a representative term for each group using a clustering algorithm that relies on semantic similarity measures. Using its default settings, REVIGO retained 70% of the terms (removing 30% of redundant ones). REVIGO also provides tree maps and other options for visualizing the subdivisions and semantic relationships for each type of GO terms. Since most of the GO terms were biological processes (BP), we used the tree maps of GO-BP terms and further modified them to summarize the GO-BP terms in each group. This analysis was performed for nominal loci associated with any mental disorders, for the Lussier study and Sharp study respectively.

### Regional Analysis

The annotation data of the 450K array also provided information on the genomic location of the DNAm loci, including whether in enhancer regions, relation to CpG-islands (i.e., Island, S shore, N shore, S shelf, N shelf, and open sea), and gene regions (i.e., gene body, 5’UTR, 3’UTR, 1st exon, TSS1500, TSS200, and intergenic). For the Lussier study, permutation tests were performed to examine whether the DNAm loci identified to be linking FASD with mental health outcomes were enriched or underrepresented in certain region. Permutation tests were performed between each pair of these three groups (ordered by increasing ranges): (1) nominal loci from the MR analyses, (2) loci input analyzed by MR, and (3) all loci tested on the 450K array. For each permutation test, we compared the percentage of loci located in a certain region, *R*, between group *A* and group *B*. If group *A* had *a* loci and group *B* had *b* loci, where *a < b*,wesampled *a* loci in the *b* loci of group *B*, computed its percentage of loci located in *R*, and repeated this random sampling process for *N* = 10,000 times. If the observed percentage of loci in group *A* located in *R*, *obs.pct_a_*, was larger or equal to the one in group *B*, *obs.pct_b_*, *p*-value would be the proportion of the *N* permutation percentages that were larger than *obs.pct_a_*, and vice versa. If *p < .*05, we concluded that the difference observed between groups was not by chance.

## RESULTS

### Validation of MR Assumptions

We confirmed the strong relevance between SNPs and DNAm loci obtained from GoDMC database by examining the maximum p-values and average absolute effect estimates (*β*) with standard deviation (SD) (**Table 2, Table S3, Table S4**), where *beta* is the percentage change of DNA methylation level for each copy of minor allele. For the Lussier study, the maximum *p*-values were 5.81×10^−9^ and 6.35×10^−20^ for *cis* and *trans* mQTL, respectively. The average of |*β*| was 0.331 for *cis* and 0.189 for *trans*. For the Sharp study, the maximum *p*-values were not larger than 1.27 × 10^−16^ for *cis* and 2.84 × 10^−23^ for *trans*. The mean of |*β*| had ranges 0.200-0.354 for *cis* and 0.096- 0.148 for *trans*.

**Table 2:** Confirmation of Relevance Assumption. Cis/Trans: whether the mQTL is *cis* or *trans*; N: the number of CpG sites available in GoDMC; |*β*|: the absolute value of the association between the SNP and DNAm; SD: standard deviation.

| Study | Pattern | Cis/Trans | N | Max P-value | Mean($\vert \beta\vert $) | SD($\vert \beta\vert $) |
| --- | --- | --- | --- | --- | --- | --- |
| Lussier | — | Cis | 168 | $5.81 \times 10^{-9}$ | 0.331 | 0.234 |
| | — | Trans | 10 | $6.35 \times 10^{-20}$ | 0.189 | 0.126 |
| Sharp | Before | Cis | 4 | $4.07 \times 10^{-21}$ | 0.276 | 0.067 |
| | Binge | Cis | 18 | $3.20 \times 10^{-26}$ | 0.354 | 0.382 |
| | Sustained | Cis | 3 | $2.74 \times 10^{-215}$ | 0.342 | 0.181 |
| | Sustained | Trans | 2 | $2.84 \times 10^{-23}$ | 0.096 | 0.012 |
| | Trimester 1 | Cis | 3 | $1.57 \times 10^{-60}$ | 0.200 | 0.063 |
| | Trimester 2/3 | Cis | 8 | $1.27 \times 10^{-16}$ | 0.244 | 0.140 |
| | Trimester 2/3 | Trans | 1 | $2.11 \times 10^{-59}$ | 0.148 | NA |

### Number of DNAm loci MR analyzed

The input DNAm loci can be lost due to missing matched SNPs in the mQTL data, which are not input into MR analysis, or missing associations between SNPs and outcomes in the GWAS data, which are lost during harmonization.

Of 161 FASD-related DNAm loci with adjusted *p < .*05 reported in the Lussier study, an average of 92 loci per outcome were associated with SNPs from the clumped mQTL data in the GoDMC database, with a maximum of 99. Most outcomes retained over 90 loci, except opioid dependence (55 loci) and suicide attempts (75 loci), which lost some SNPs during harmonization (**Figure 2, Table S15**).

**Figure 2:**
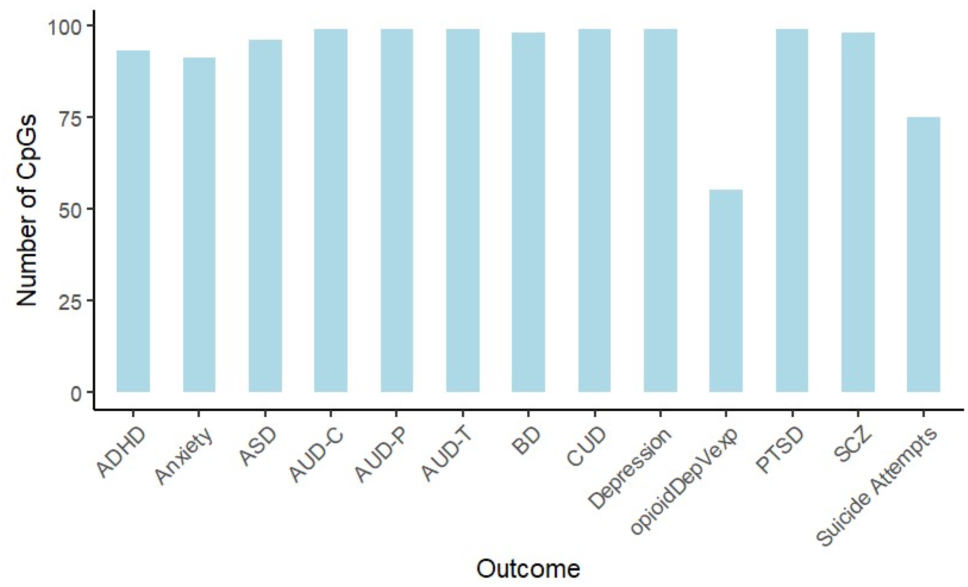
Number of DNAm loci analyzed for each outcome from the Lussier study. OpioidDepVexp: opioid dependence (versus exposure).

Of 53 DNAm loci with unadjusted *p <* 1 × 10^−5^ in the Sharp study, 25 were associated with SNPs from the clumped mQTL data in the GoDMC database, some of which were dropped out during harmonization due to lack of matched data in the outcome GWAS. Binge drinking had the most MR-analyzed DNAm loci (at most 13 loci), followed by drinking during trimesters 2 or 3 (at most 6 loci) (**Figure 3**). Most of the outcomes analyzed all of the available loci, but opioid dependence again lost the most loci during harmonization (**Figure 4, Table S16**), similar to the condition in the analysis for the Lussier study.

**Figure 3:**
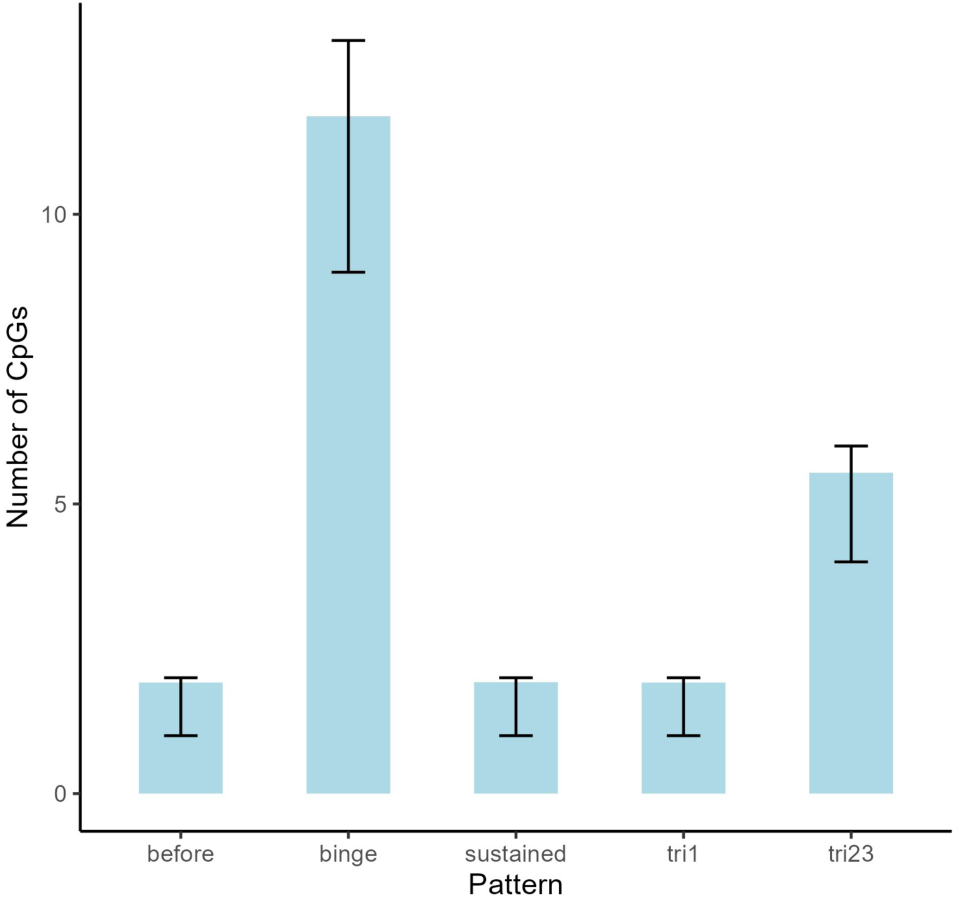
Number of MR-analyzed DNAm loci per drinking pattern from the Sharp study. The height of each bar shows the mean number. Each error bar indicates the range from minimum to maximum across all outcomes. tri1: trimester 1; tri23: trimester 2 or 3

**Figure 4:**
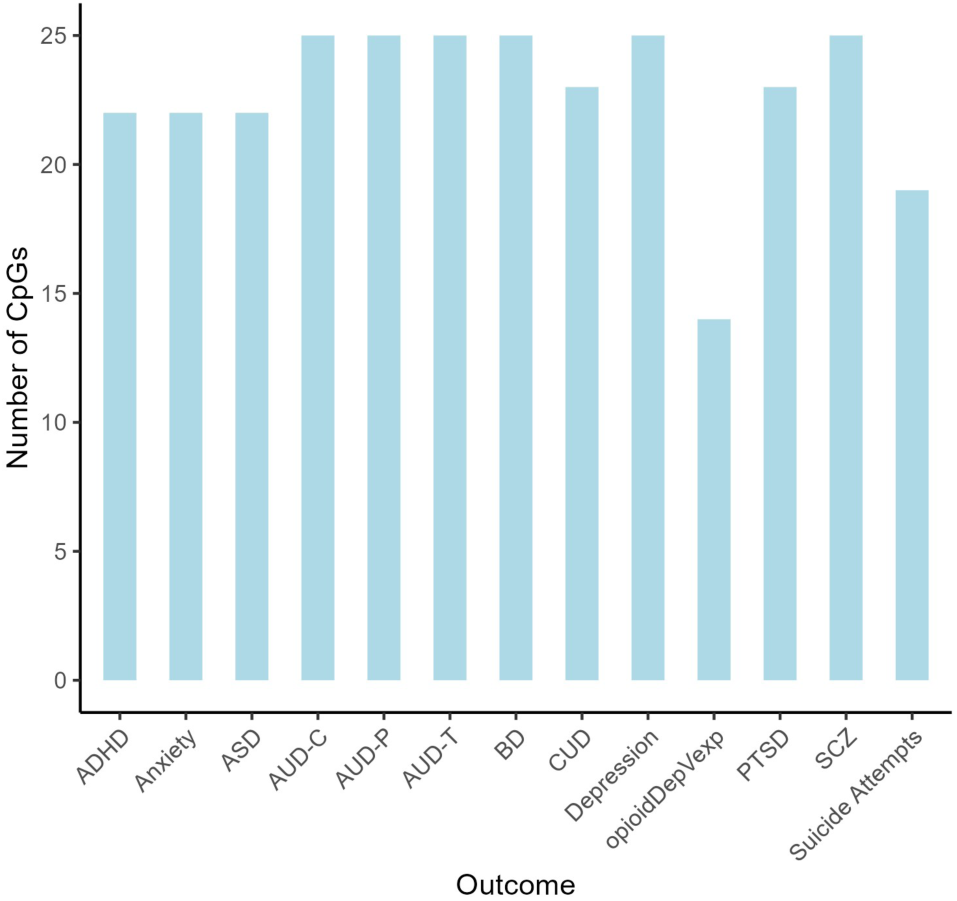
Number of MR-analyzed DNAm loci per outcome, summed over all patterns, from the Sharp study.

### Associations between PAE-related DNAm loci and Adverse Mental Health Outcomes

In general, PAE have negative impact on offspring health and DNAm is assumed to increase risk of adverse mental health outcomes. However, DNAm may also adapt to suppress the negative impact rather than solely increasing the risk [44]. Thus, we assessed whether the role of PAE-related DNAm was to increase or suppress the risk of mental illness.

To investigate the role of DNAm in linking adversity to mental disorders, we computed an adjusted MR estimate (*adj.beta*) by multiplying the unadjusted MR estimate (*b*) by the sign of PAE- DNAm association (*delta.beta*) established previously in the EWAS (**Table S13, Table S14**). *delta.beta* was extracted from the *X.beta* column in **Table S2**. *adj.beta >* 0 means that PAE raises the risk of mental illness through altering DNAm level at the CpG site, while *adj.beta <* 0 means PAE decreases the risk (**Table 3**):

**Table 3:** Direction of effects among PAE, DNAm, and mental disorder risk. *delta.beta*: estimate of the associations between PAE or FASD and DNAm from the EWAS (i.e. Lussier study and Sharp study). *b*: raw estimate of the associations between DNAm and risk of mental disorders from MR. *adj.beta*:

| **PAE ~ DNAm**<br>**(delta.beta)** | **DNAm ~ Mental**<br>**Disorder (b)** | **PAE ~ Mental**<br>**Disorder (adj.beta)** |
| --- | --- | --- |
| + | + | + |
| - | - | + |
| + | - | - |
| - | + | - |

In the Lussier Study, a total of 108 associations between DNAm and mental health outcomes reached nominal significance (*p < .*05, **Table S13**). Among these, 36 had *p < .*01, and 19 remained significant after Benjamini–Hochberg correction (*FDR < .*05). None of these associations failed the heterogeneity test or the pleiotropy test, while 2 associations failed the leave-one-out test (**Figure S1, S4; Table S7-S9**). After excluding these 2 associations, 106 associations with *p < .*05 remained, 36 with *p < .*01, and 19 with *FDR < .*05 (**Figure 5**). The nominal associations that passed the sensitivity tests involved a total of 55 DNAm loci (*p < .*05). Specifically, depression had the most associations with DNAm (16 loci), followed by SCZ (14 loci), BD (13 loci) and ASD (9 loci). Among the nominal loci, cg14228272 had the most associations. The strongest association was between cg26112661 and SCZ (*adj.beta* = 1.04*, FDR* = 8.31 × 10^−18^). The association between cg26112661 and SCZ was *b* = −1.04. Given the negative association between FASD and cg26112661 (*delta.beta* = −0.04 *<* 0), the adjusted estimate suggests that PAE is positively related to SCZ through this CpG site. In other words, PAE-associated DNAm change at cg26112661 may increase risk for SCZ.

**Figure 5:**
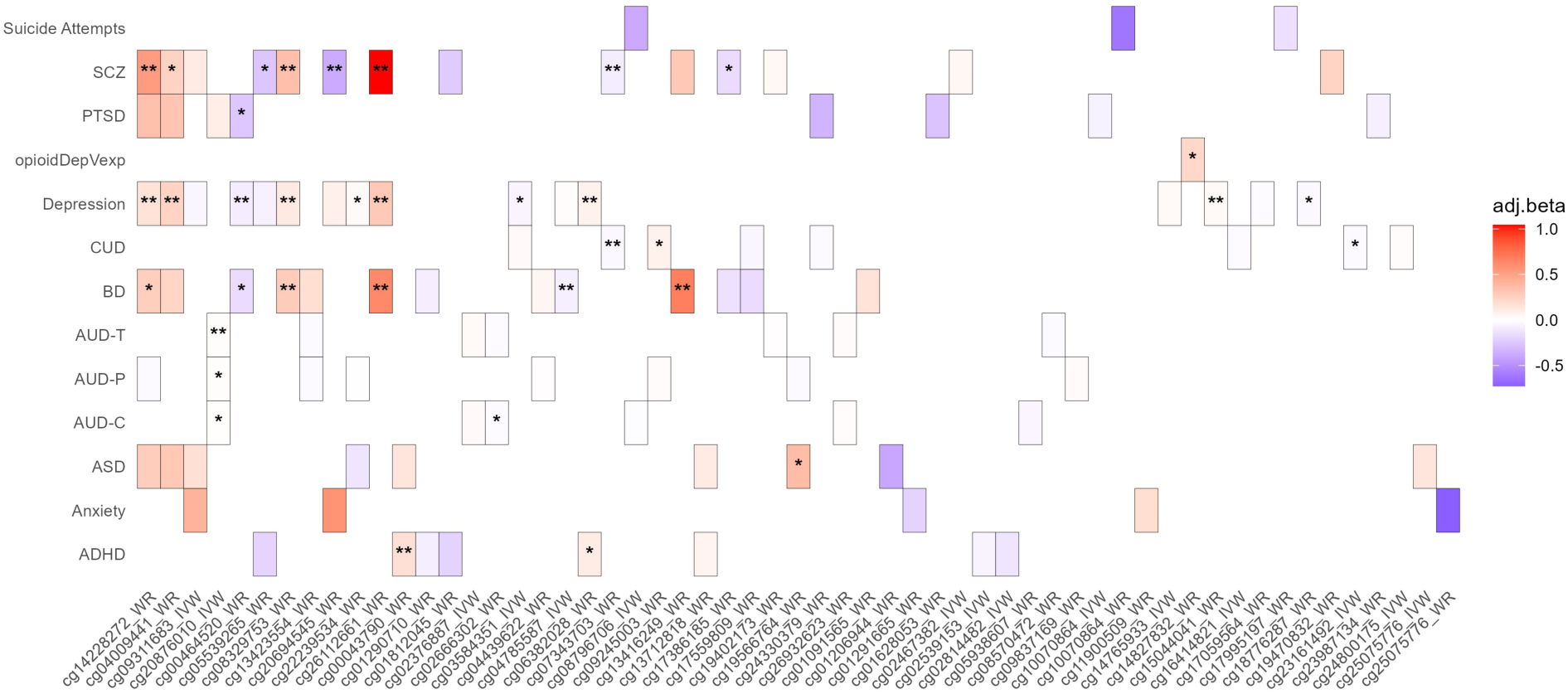
Heatmap of DNAm-mental health outcome associations with *p < .*05, excluding those failing sensitivity test(s). *Note:* ∗*p < .*01, ∗∗*FDR < .*05. Blue for *adj.beta <* 0, corresponding to risk-suppressing DNAm loci; red for *adj.beta >* 0, corresponding to riskincreasing DNAm loci. The x-axis are CpG labels consisted with two parts, CpG ID and MR method. IVW: inverse variance weighting method; WR: Wald Ratio method. The CpG labels are ordered by the number of outcomes they are associated with. The y-axis are outcomes. OpioidDepVexp: opioid dependence (versus exposure).

For the Sharp Study, a total of 28 DNAm–mental health associations reached nominal significance at *p < .*05 (**Table S14**). Of these, 12 met a more stringent threshold of *p < .*01, and 8 remained significant after multiple testing correction (*FDR < .*05) (**Figure 6**). None of the associations failed any sensitivity tests, but this result can be owing to insufficient number of SNPs available to conduct the tests ((**Figure S3; Table S10-S12**)). Binge drinking had the most associations, which is natural since it had the most input loci. Schizophrenia and bipolar disorder had the most associations with PAE-related DNAm loci (5 for each). **Figure 7** categorized the effect of each pattern of drinking on offspring mental health outcome into 3 classes, risk-increasing, risk-suppressing, and mixed effect. Binge drinking can increase risk of anxiety, AUD-C, BD and CUD, decrease risk of AUD-P, ASD, depression, and PTSD, and had a mixed effect on SCZ, through DNAm. Drinking during trimester 2 or 3 had protective effects on ADHD, BD, depression, SCZ, and suicide attempts, with slight risk-increasing effect on AUD-T and AUD-C. Other patterns had fewer input loci and thus had fewer associations, with only 1 or 2 loci identified. Through DNAm changes, alcohol drinking before pregnancy may increase risk of BD, sustained drinking may increase risks of SCZ and BD, while drinking during trimester 1 had protective effects on PTSD, CUD, and AUD-C.

**Figure 6:**
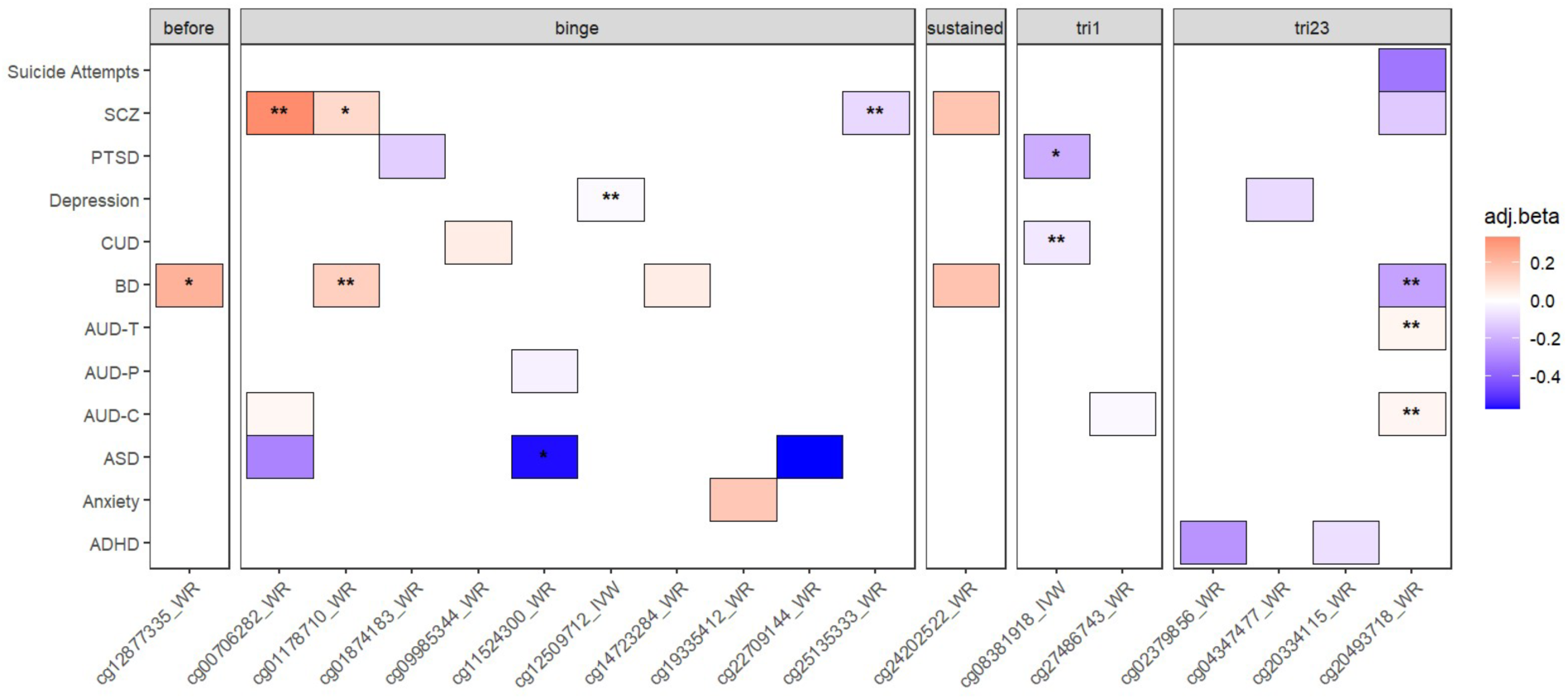
Heatmap of DNAm-mental health outcome associations with *p < .*05. *Note:* ∗*p < .*01, ∗∗*FDR < .*05. Blue for *adj.beta <* 0, corresponding to risk-suppressing DNAm loci; red for *adj.beta >* 0, corresponding to risk-increasing DNAm loci. The x-axis are CpG labels consisted with two parts, CpG ID and MR method. IVW: inverse variance weighting method; WR: Wald Ratio method. The y-axis are outcomes. OpioidDepVexp: opioid dependence (versus exposure).

**Figure 7:**
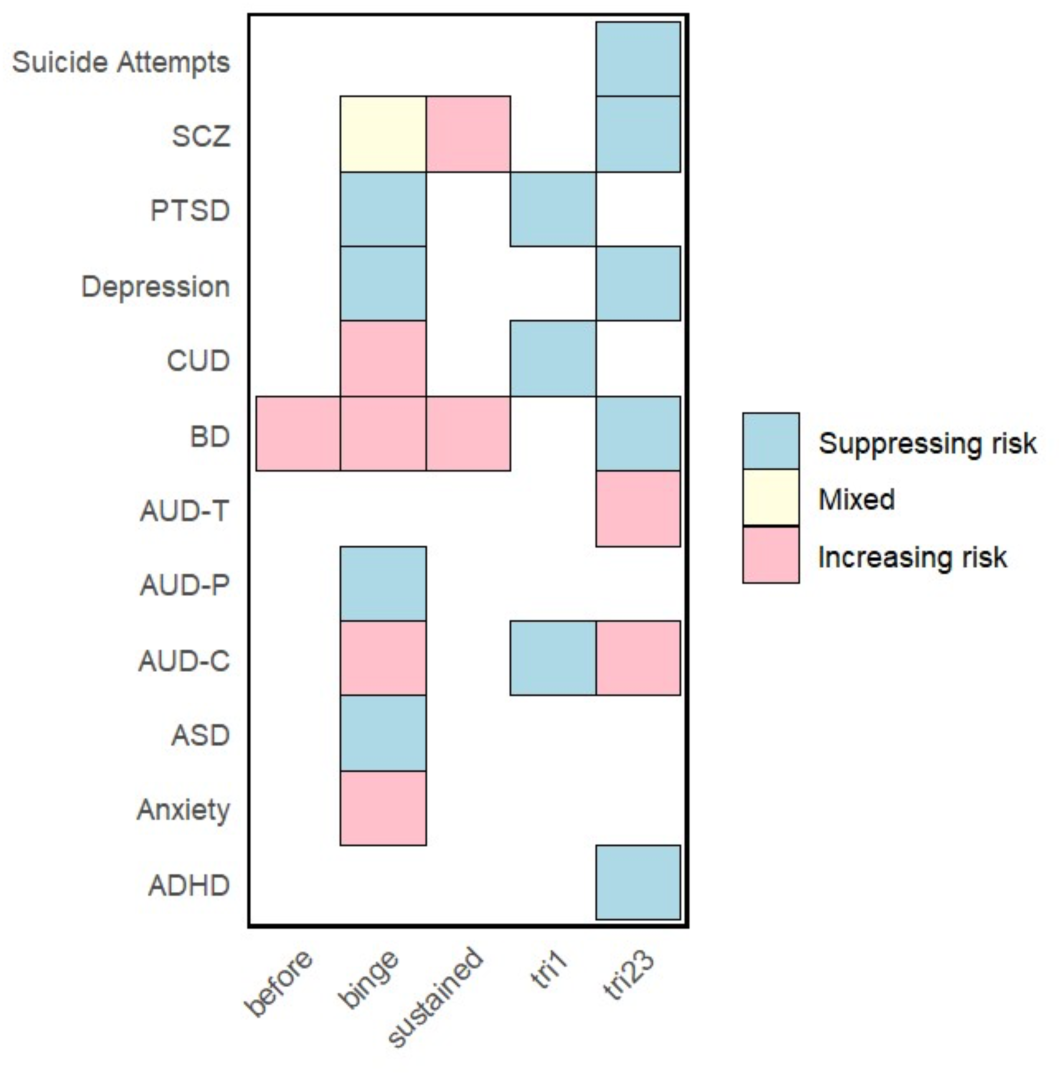
Risk Effects of different patterns of PAE for each adverse mental health outcome. Pink: all CpG sites had risk increasing effect on the outcome. Blue: all CpG sites had risk suppressing effect on the outcome. Yellow: some CpG sites increased risk, while the others decreased risk.

Notably, 12 DNA methylation loci among the nominal loci that passed the sensitivity tests were identified as expression quantitative trait methylation (eQTM) loci (**Table S13**), whereas none of the nominal associations in the Sharp Study overlapped with known eQTM loci. The associations involving these loci should be interpreted with caution, since DNAm may not be the only pathway through which SNPs impact on mental health outcomes. Moreover, for the Lussier study, the association between cg04009441 and depression could be invalid, since the SNP rs12521969 associated with this CpG site had known association with depression, according to a GWAS on depression including *>* 1.3 million individuals (*p* = 2 × 10^−15^, **Table S3**) [37].

The two EWAS revealed differing ratios of risk-increasing and risk-suppressing DNAm loci associated with each outcome. Only loci with *p < .*05 and passing all sensitivity tests were included in this comparison (**Figure 8**). In particular, Opioid dependence versus exposure in the Sharp study had no association with DNAm loci with *p < .*05.

**Figure 8:**
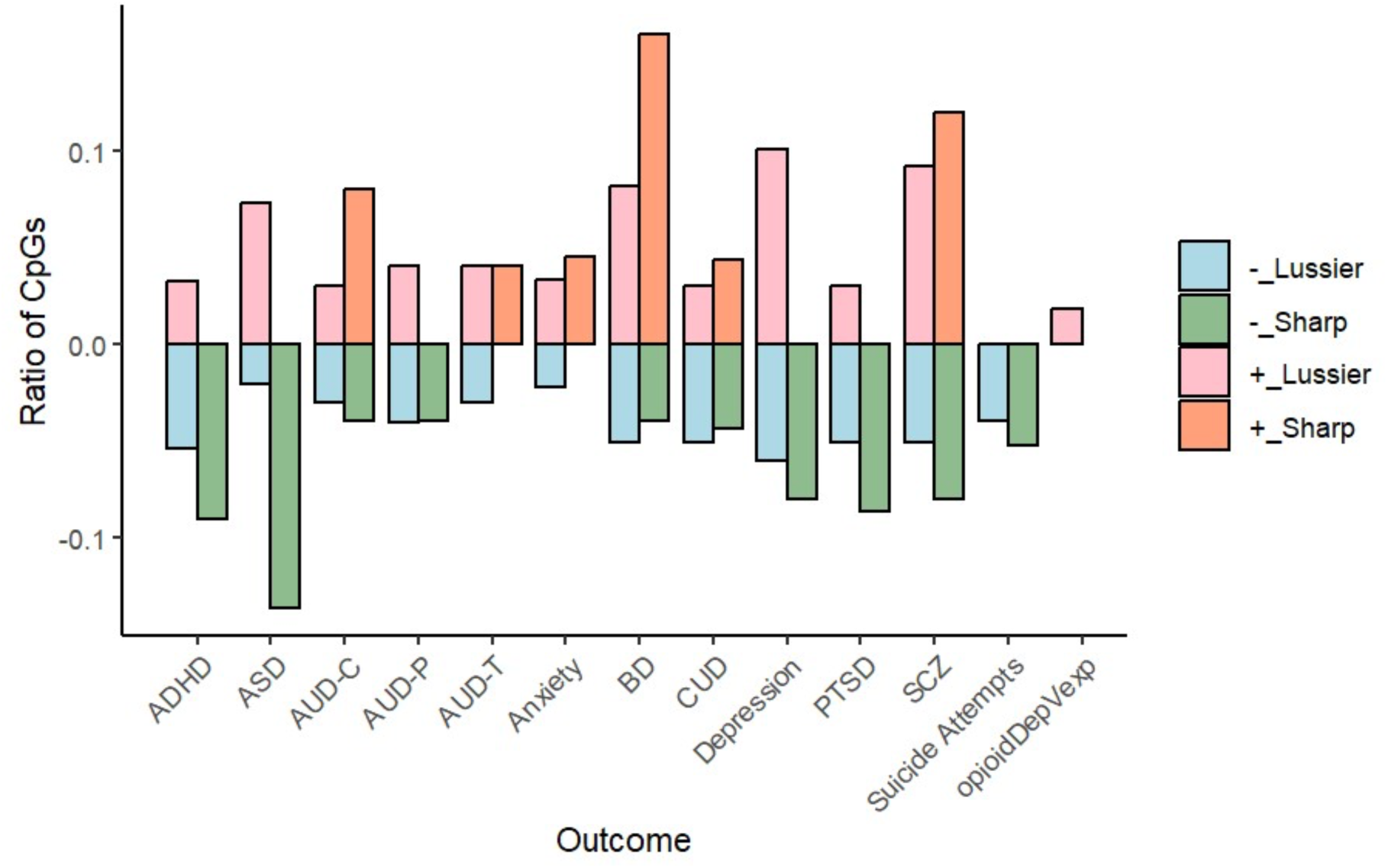
Ratios of risk-increasing and risk-suppressing DNAm loci with *p < .*05 and that passed the sensitivity tests, for each outcome and study.

### Functional Enrichment Analysis

Of the 55 DNAm loci from the Lussier study associated with mental illness, 31 had risk-increasing effects on at least one mental health outcome, while 33 had risk-suppressing effects. The risk-increasing DNAm loci were relevant to cell fate specification, heart development, response to ions, and axonal protein transport (**Figure 9**). By contrast, the risk-suppressing loci were involved in carbohydrate and nucleotide catabolism, protein translation, membrane organization, immune signaling, response to ions, and dopaminergic synaptic transmission (**Figure 10**).

**Figure 9:**
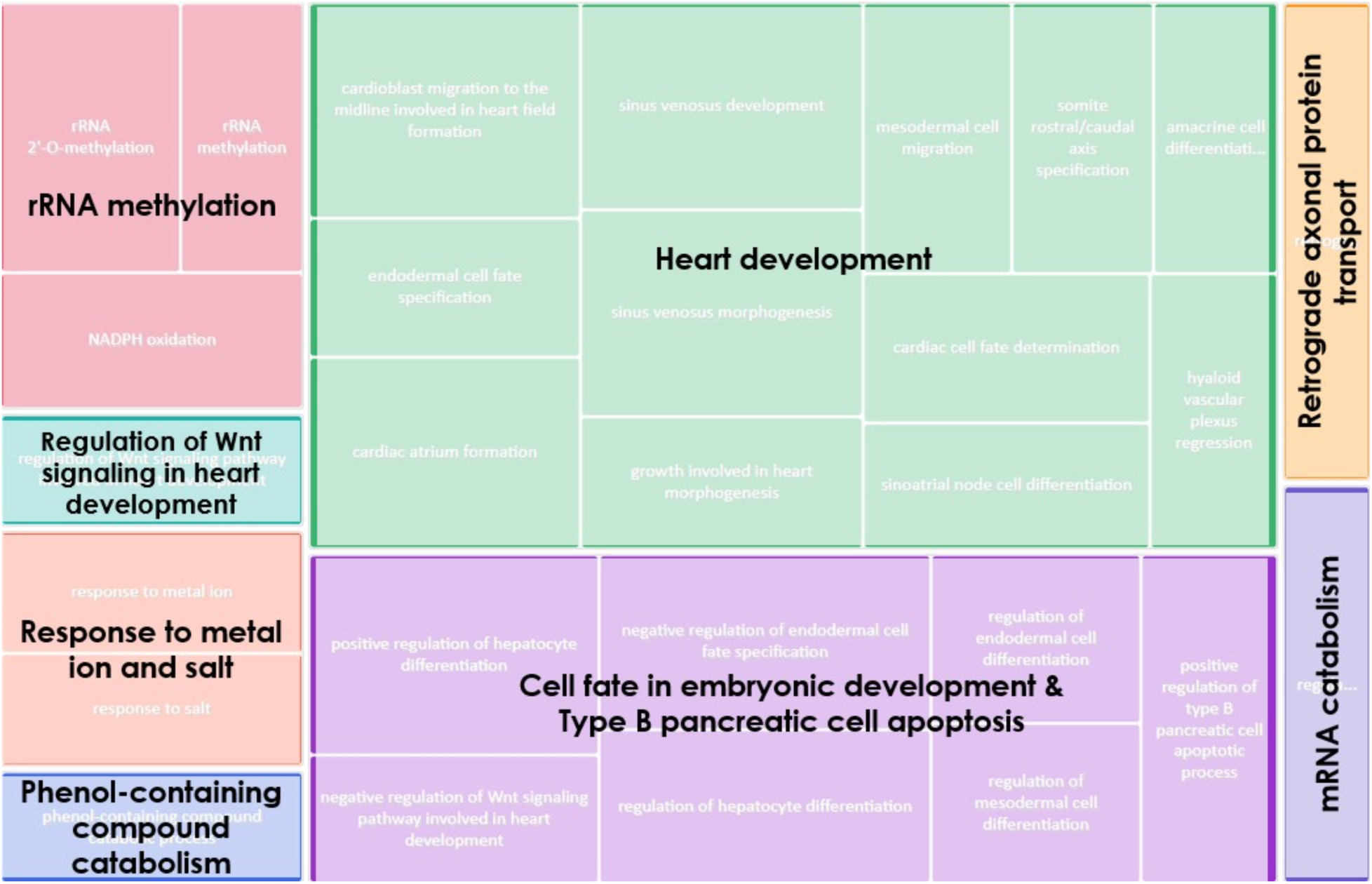
Biological Processes (BPs) involved by the risk-increasing DNAm loci in the Lussier study. The top 100 Gene Ontology biological process (GO-BP) terms, based on p-values from the gometh() function applied to 26 nominally significant CpG sites with *adj.beta >* 0, were submitted to REVIGO for redundancy reduction and clustering. REVIGO then generated this tree map, where GO-BP terms were grouped by semantic similarity, with terms in the same group sharing the same color. Group-level biological themes were manually summarized and overlaid on the corresponding areas of the tree map.

**Figure 10:**
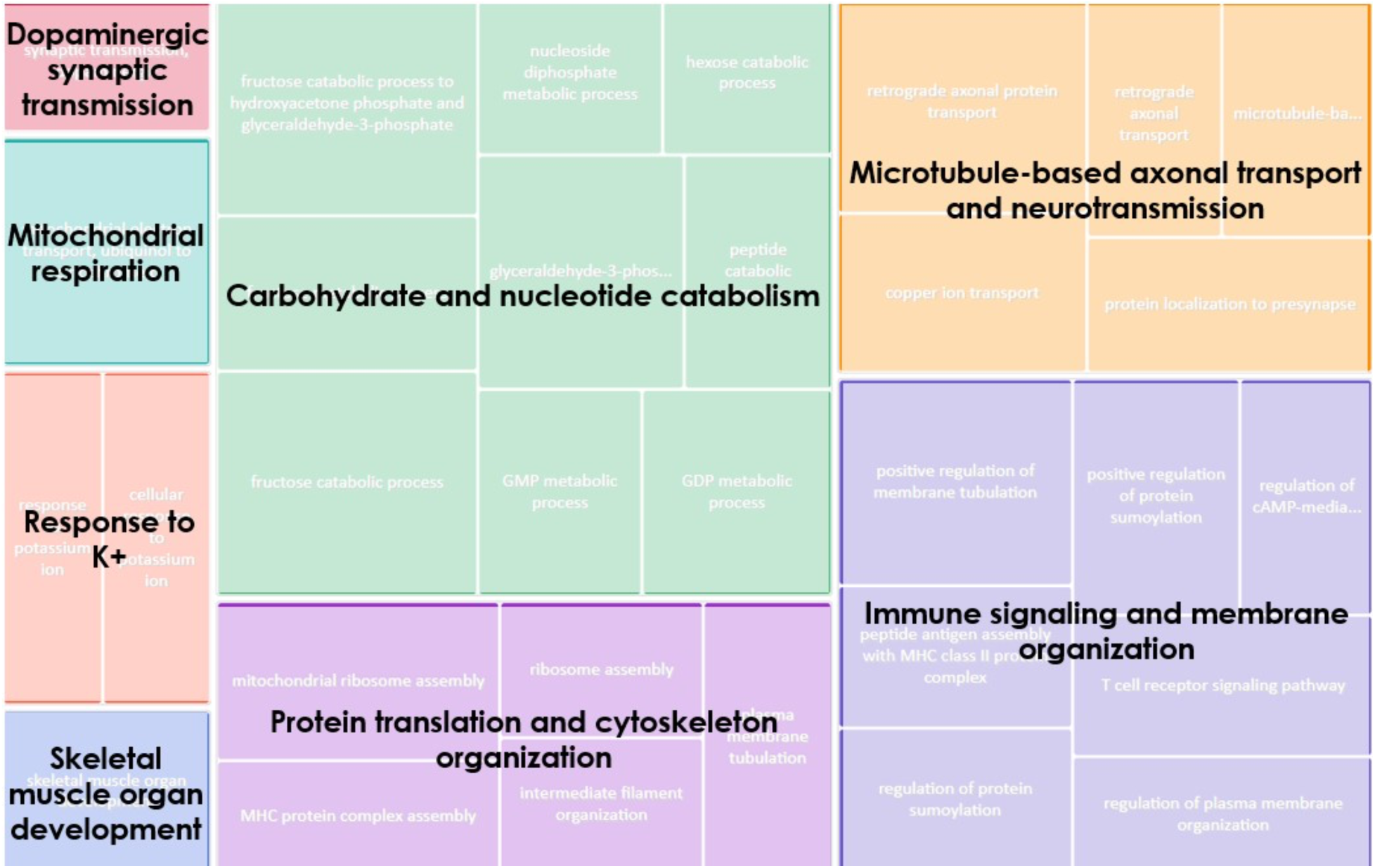
Biological processes involved by the risk-suppressing DNAm loci in the Lussier study. The top 100 Gene Ontology biological process (GO-BP) terms, based on p-values from the gometh() function applied to 18 nominally significant CpG sites with *adj.beta <* 0, were submitted to REVIGO for redundancy reduction and clustering. REVIGO then generated this tree map, where GO-BP terms are grouped by semantic similarity, with terms in the same group sharing the same color. Group-level biological themes were manually summarized and overlaid on the corresponding areas of the tree map.

**Figure 11:**
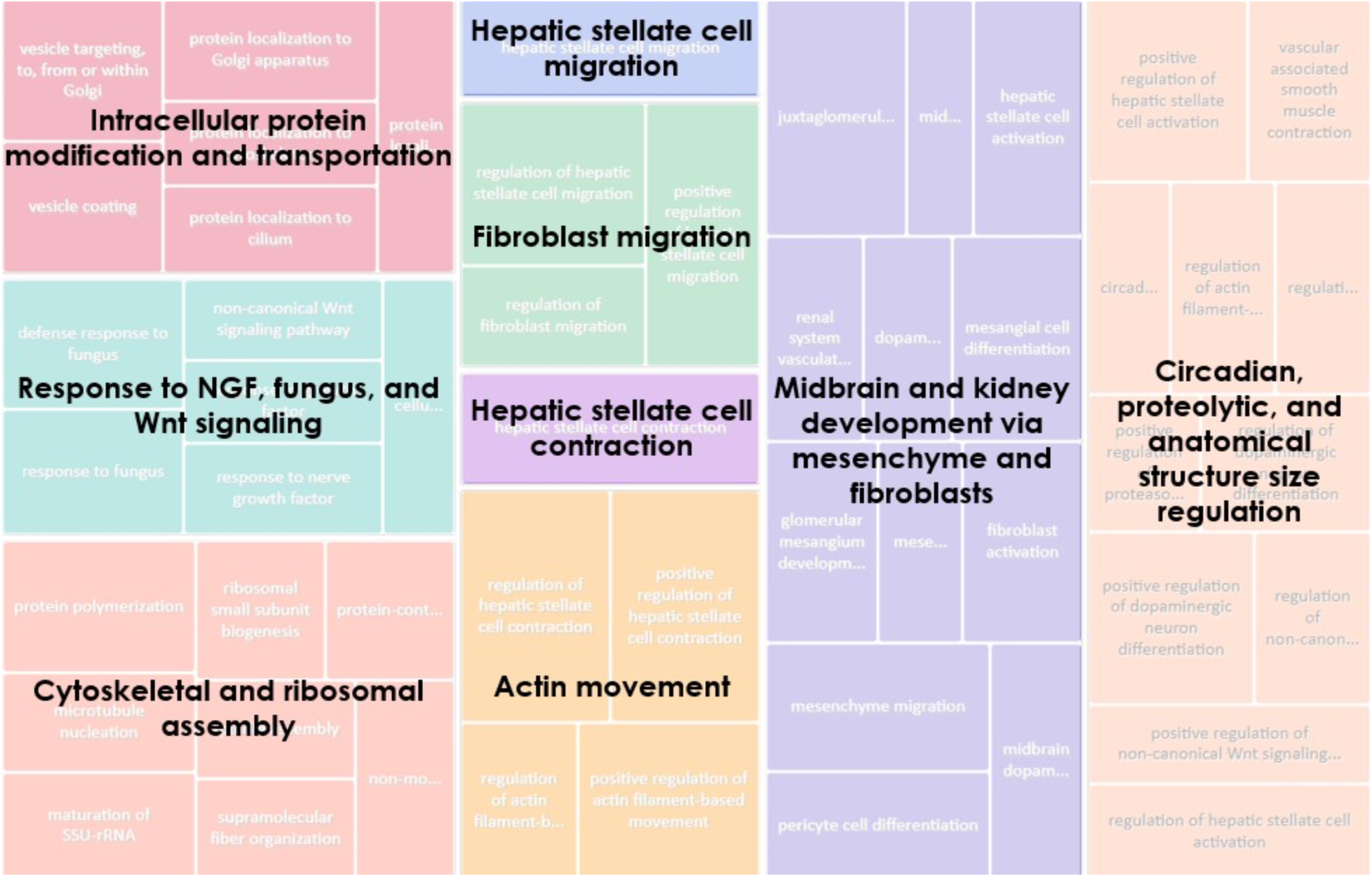
Biological Processes involved by the risk-increasing DNAm loci in the Sharp study. The top 100 Gene Ontology biological process (GO-BP) terms, based on p-values from the gometh() function applied to 8 nominally significant CpG sites with *adj.beta >* 0, were submitted to REVIGO for redundancy reduction and clustering. REVIGO then generated this tree map, where GO-BP terms are grouped by semantic similarity, with terms in the same group sharing the same color. Group-level biological themes were manually summarized and overlaid on the corresponding areas of the tree map.

**Figure 12:**
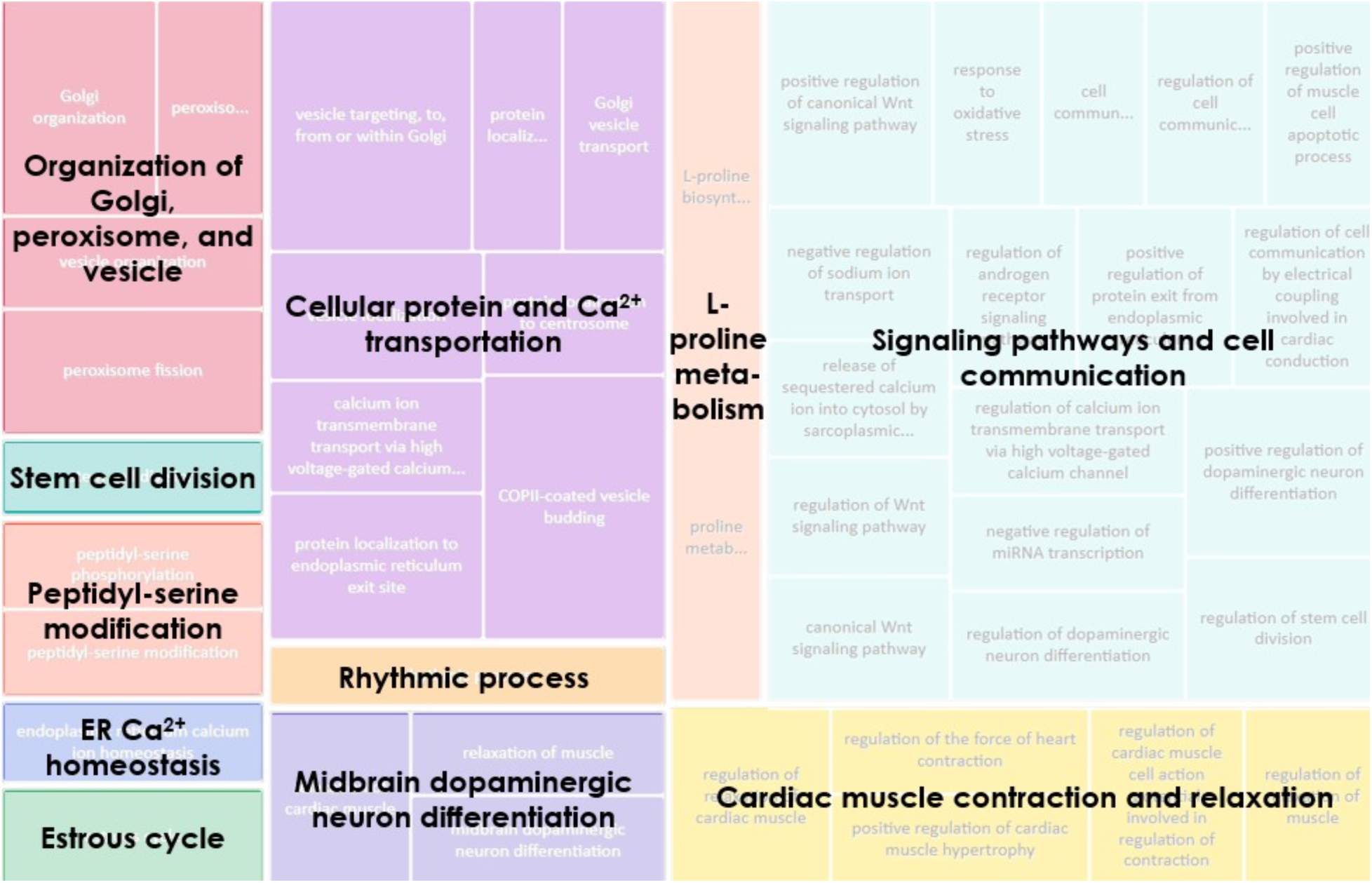
Biological Processes involved by the risk-suppressing DNAm loci in the Sharp study. The top 100 Gene Ontology biological process (GO-BP) terms, based on p-values from the gometh() function applied to 12 nominally significant CpG sites with *adj.beta <* 0, were submitted to REVIGO for redundancy reduction and clustering. REVIGO then generated this tree map, where GO-BP terms are grouped by semantic similarity, with terms in the same group sharing the same color. Group-level biological themes were manually summarized and overlaid on the corresponding areas of the tree map.

Of the 18 DNAm loci from the Sharp study associated with mental disorders, 8 had risk-increasing effects and 12 had risk-suppressing effects. CpG sites involving post-translation modification and transportation of protein, as well as dopaminergic neuron differentiation, had both risk-increasing and risk-suppressing effects. Specifically, risk-increasing loci involved midbrain and kidney development, response to nerve growth factor (NGF), fungus, and non-canonical Wnt signaling, hepatic stellate cell contraction and migration driven by actin movement, and regulation of circadian gene expression, protein catabolism, and anatomical structure. By contrast, risk-suppressing DNAm loci involved mainly in stimulus response involving cardiac muscle and calcium ion (Ca2+) transmembrane transport.

### Regional Enrichment Analysis

The 99 MR-analyzed FASD-related DNAm loci from the Lussier study were enriched in enhancer regions compared to all the 450K loci (*p* = .021, **Figure 13d**). The nominal loci further associated with mental health showed a similar pattern to these loci, showing non-significant enrichment in enhancers compared to the 450K loci (*p* = 0.264, **Figure 13b**). From the perspective of gene region, the MR-analyzed FASD-related loci were unlikely to be located within 200 base pairs upstream to transcription start sites (TSS200), compared to all 450K loci (*p* = .001, **Figure 14d**). Moreover, none of the nominal loci associated with mental health located in TSS200 (**Figure 14a**). As for relation to CpG islands, the MR-analyzed FASD-related DNAm loci were more likely to appear in S shores, which is within 2kb downstream to CpG Islands, compared to the 450K loci (*p* = 0.044; **Figure 15d**). The regional analysis was not performed for the Sharp study due to the limited number of nominal loci, which may lead to unstable p-value estimates.

**Figure 13:**
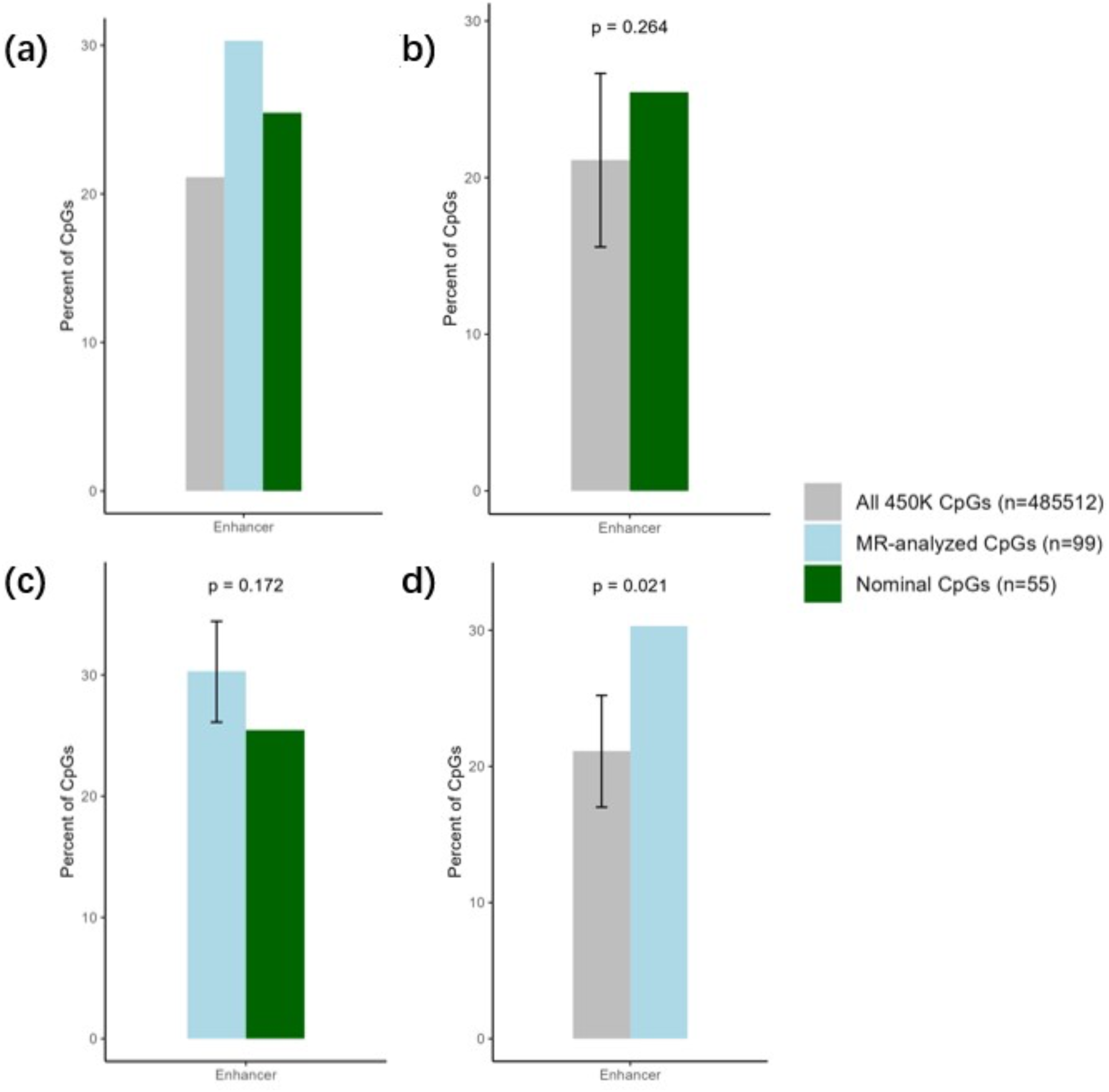
Permutation test results for whether the percentage of DNAm loci located in enhancer differed between groups. (a) % DNAm loci in enhancer regions. (b) Permutation test between nominal loci and all 450K-tested loci. (c) Permutation test between nominal loci and MR-analyzed loci. (d) Permutation test between MR-analyzed loci and all 450K loci. In (b)(c)(d), *p < .*05 indicates that the percentage of DNAm loci located in enhancer differed between groups not by chance. The error bars shows the range *mean* ± *sd*.

**Figure 14:**
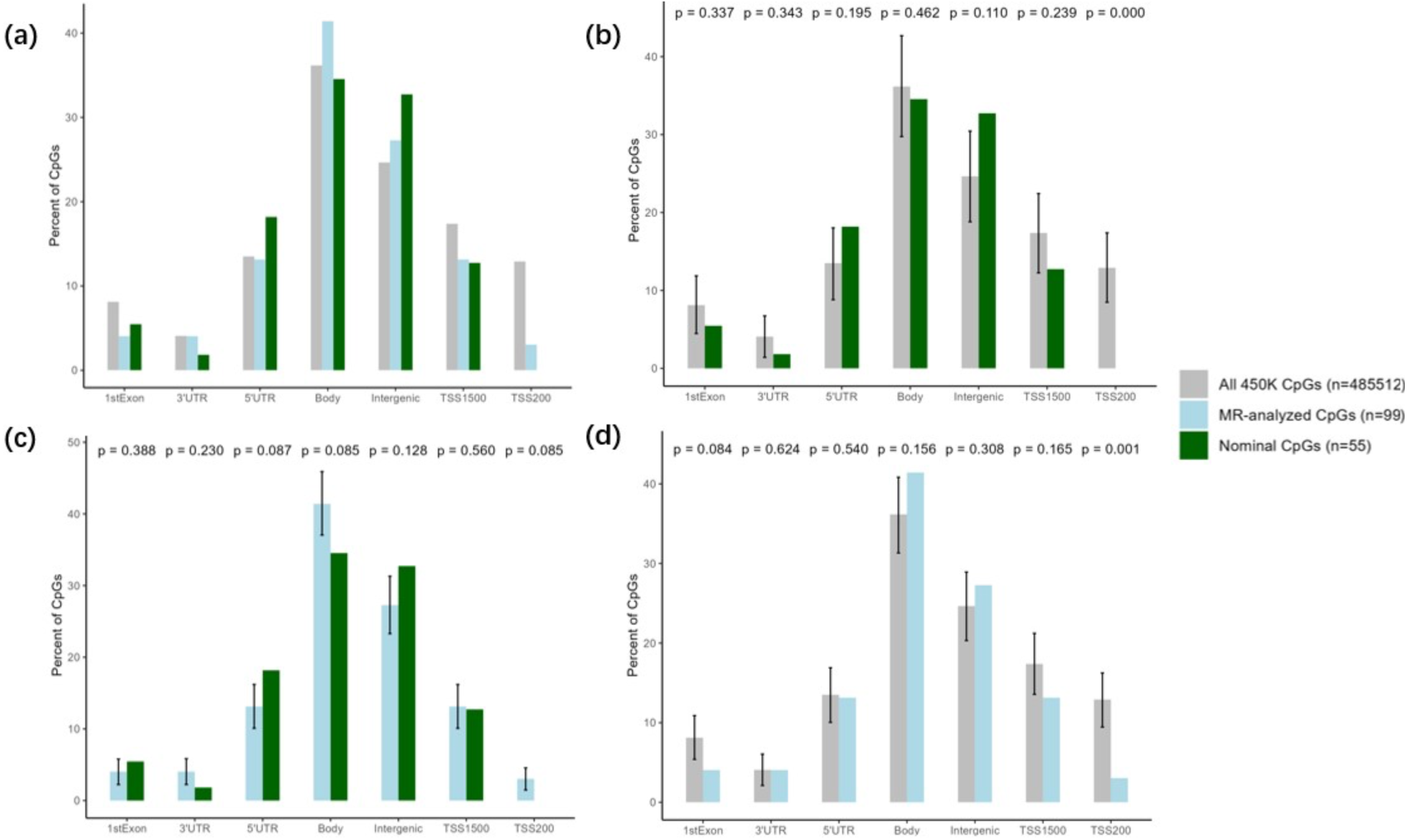
Permutation test results for whether the percentage of DNAm loci located in genomic regions differed between groups. (a) % DNAm loci in different gene regions. (b) Permutation test between nominal loci and all 450K-tested loci. (c) Permutation test between nominal loci and MR-analyzed loci. (d) Permutation test between MR-analyzed loci and all 450K-tested loci. In (b)(c)(d), *p < .*05 indicates that the percentage of DNAm loci located in the gene region differed between groups not by chance. The error bars shows the range *mean* ± *sd*. Body: gene body; 1stExon: the first exon; 5’UTR: 5’ untranslated region; 3’UTR: 3’ untranslated region; TSS1500: within 1500 bp upstream to the transcription start site; TSS200: within 200 bp upstream to the transcription start site; Intergenic: region between genes. 5’UTR, TSS1500, and TSS200 are commonly considered as promoter regions. Together with 1stExon, these regions are where DNAm is generally associated with transcriptional repression.

**Figure 15:**
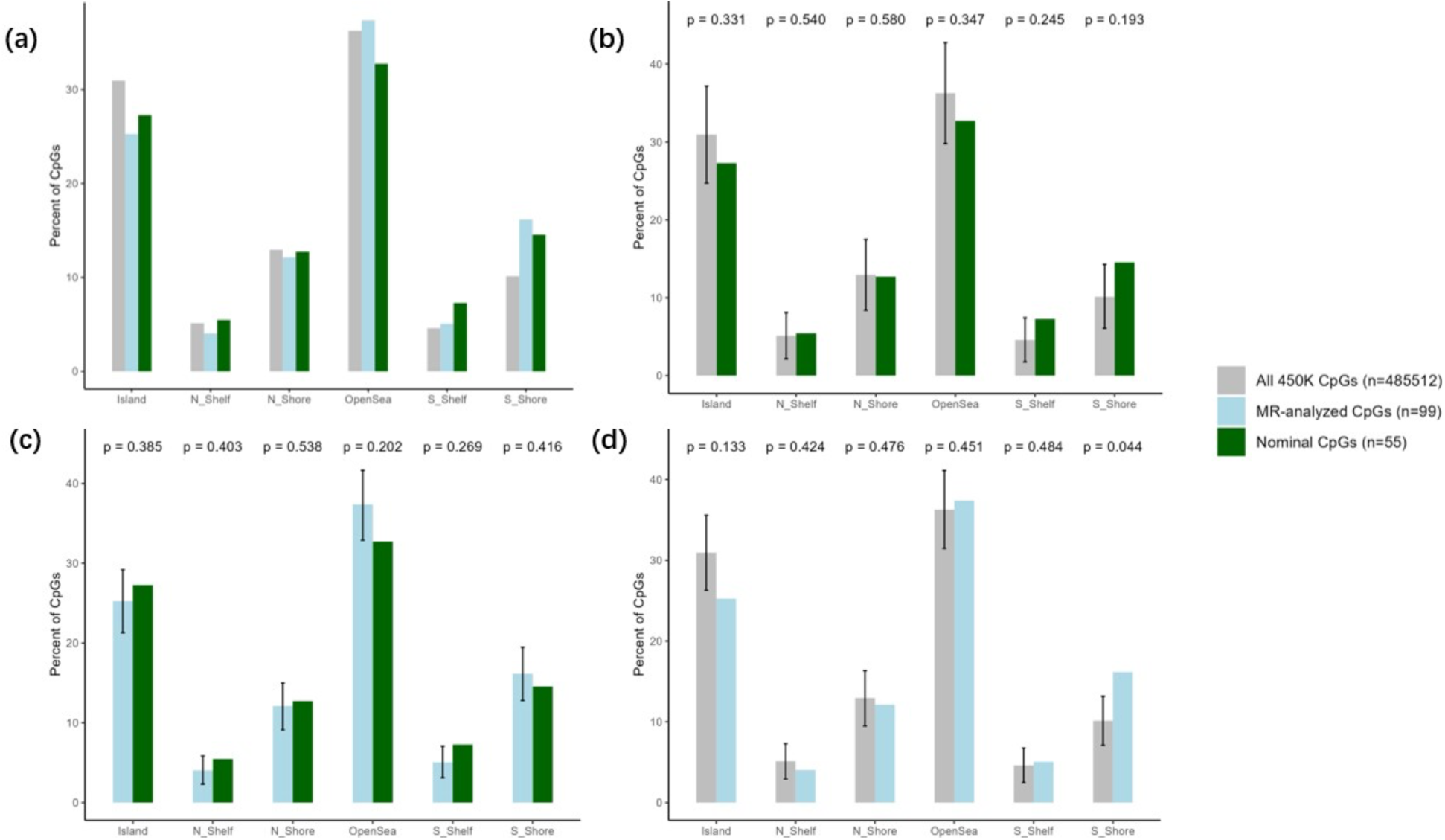
Permutation test results for whether the percentage of DNAm loci’s relations to CpG Islands differed between groups. (a) % DNAm loci in different relations to CpG islands. (b) Permutation test between nominal loci and all 450K-tested loci. (c) Permutation test between nominal loci and MR-analyzed loci. (d) Permutation test between MR-analyzed loci and all 450K-tested loci. In (b)(c)(d), *p < .*05 indicates that the percentage of DNAm loci with such relation to CpG islands differed between groups not by chance. The error bars shows the range *mean* ± *sd*. Island: CG Island, a region with high cytosine- and guaning content; N Shore/S Shore: upstream (5’/North) and downstream (3’/South) shore regions up to 2kbp-wide regions flanking the CG islands; N Shelf/S Shelf : The upstream (5’/North) and downstream (3’/South) shelf regions 2-4kbp-wide regions flanking CG Islands and their shores; OpenSea: any regions occurring between CG islands.

## DISCUSSION

This study identified DNAm loci from two previous EWAS studies that may link PAE to adverse mental health outcomes across the life course. Loci from both studies showed that SCZ and BD had the most associated DNAm loci, followed by depression and ASD, suggesting that the effects of PAE on the risk for these mental disorders may be more likely to occur through epigenetic mechanisms. Both the Lussier study and the Sharp study had slightly more risk-suppressing loci than risk-increasing ones. Nonetheless, both studies had some loci with mixed effects, influencing different adverse mental health outcomes in different directions.

Analyses stratified by drinking pattern revealed a complex and nuanced landscape of how distinct prenatal drinking patterns may influence later mental health outcomes through epigenetic modifications. Rather than a uniform risk effect, different exposure patterns appeared to confer heterogeneous and sometimes opposing effects on various psychiatric phenotypes. Such heterogeneity may arise from: (1) Pattern-specific effects of PAE on DNAm, as observed in the Sharp study, where different PAE patterns were associated with distinct sets of DNAm loci showing both positive and negative associations. The direction of these associations may depend on the genomic location of the DNAm loci, which leads to up- or down-regulated gene expression. Plus, the role that DNAm plays may be complex. Not only can it be risk-increasing but also buffering the negative impact. (2) Pattern-specific effects of PAE on mental health, as found in previous studies, first-trimester moderate alcohol exposure is associated with increased risk of anxiety or depression in children [49], first-trimester heavy alcohol exposure is associated with offspring alcohol use [7], whereas other combinations of dose and timing of PAE did not show significant associations with offspring mental illness in these studies. This variability can possibly be explained by timing-specific developmental vulnerabilities of brain, where different gestational stages correspond to the maturation of different brain regions, making some periods more sensitive to epigenetic disruption. In other words, the impact of PAE on mental health is shaped by the specific pattern and timing of exposure, emphasizing the biological complexity underlying FASD-related outcomes.

Functional enrichment analyses provided additional insights into the potential biological mechanisms that link PAE to mental illness. DNAm loci identified across both studies were involved in signaling pathways (via Wnt pathways, ions, protein, and other factors), cell differentiation for development, and cytoskeletal structure organization. These findings strengthened the biological plausibility of the MR results, suggesting that at least some of the identified DNAm loci may play roles in pathways through which PAE contributes to neurodevelopmental and psychiatric outcomes. This is consistent with the deficits observed in FASD, which impact memory, behavior, and brain structure and function [50]. For the neural system, considered to be most related to mental health, the DNAm loci from the Lussier study are involved in axonal protein transportation, protein translation and localization to presynapse, dopaminergic synpatic transmission, and response to ions. Likewise, the loci from the Sharp study are involved in dopaminergic neuron differentiation and cell communication through ion transport. These findings are also consistent with prior literature and may provide new insights. For example, PAE can dysregulate genes associated with regulation of synaptic transmission, transport, and synaptic vesicle cycle in the mouse brain, which further impact cognition and behavior [51]. Animal studies have also demonstrated that PAE can inhibit the lesion-induced dopaminergic synapse responsiveness in the olfactory tubercle [52], and early-gestation moderate PAE can blunt the dopaminergic function [53]. These findings revealed the critical impact of PAE on fetal development, especially for pathways involved in the neural system. The regional analysis for the Lussier study showed that the FASD-related DNAm loci are more likely to locate in enhancer regions and S-shore of CpG islands, but are underrepresented in TSS200 regions, indicating a potential role for distal regulatory elements. Disruptions in enhancer activity have been linked to various brain diseases, highlighting their importance in maintaining neural health [54]. Methylation at CpG island shore regions can regulate genes involved in stress response [55]. Nevertheless, the findings of the regional analysis remained exploratory due to the limited number of loci and annotations tested.

This study had several strengths. The two-sample MR analysis was conducted with caution to validate its three assumption. We used clumped data from the GoDMC database to ensure that LD had been adjusted for from the original studies, instead of using a general approach to clump the results after MR analysis. GoDMC is the largest study of mQTLs, encompassing data from more than 30,000 participants, which ensured that we selected robust IVs for our MR analyses. For associations between SNPs and outcomes, we used summary statistics from publicly available GWAS with large sample sizes, enhancing the statistical power of our estimation. Additionally, we investigated drinking pattern-specific (i.e., binge, sustained, and trimester-specific) associations between DNAm and mental health outcomes, offering a more granular view of how different PAE patterns confer differential risk for mental disorders through epigenetic regulation.

Nonetheless, several limitations should be acknowledged. First, the IVW method relies strictly on the exclusion restriction assumption, which may not be necessarily upheld even though we had performed the sensitivity tests. As mentioned before, skipped tests due to insufficient SNPs, eQTM DNAm loci, and eQTL SNP with known effect on outcome, posed threats to this assumption. Thus, the results involving eQTM DNAm loci or eQTL SNP of the Lussier study (**Table S13**), as well as the results with missing test results, especially for the Sharp study (**Table S10-S12**), should be interpreted with caution. Second, bidirectional MR was not feasible due to data limitations, leaving open the possibility of reverse causality. However, this limitation is mitigated by the fact that the DNAm samples were collected at birth (Sharp) or in children and adolescents (Lussier), as many of the analyzed mental disorders emerge later in life. Third, tissue mismatch between exposure data in the Lussier study (i.e., buccal cells) and mQTL sources (i.e., blood samples) may also limit biological interpretability and the robustness of the associations. Fourth, the cohorts used were predominantly composed of individuals of Caucasian ancestry, which helped reduce potential bias due to population stratification, but limited the potential to generalize our findings to more diverse populations. Fifth, cutoffs using unadjusted *p*-value may be less robust due to multiple testing effect, so we provided adjusted *p*-values in the full results (**Table S5-S6**) and denoted them in the graphs. Last but not least, the mental disorders with the highest number of associated loci, namely SCZ, BD, depression, and ASD, are known to have substantial genetic contributions [45, 46, 47, 48]. Therefore, the observed associations may be influenced by genetic confounding. In other words, individuals with a higher genetic predisposition to these disorders may be more likely to engage in alcohol consumption and have children with PAE.

Future research should seek to replicate and expand upon these findings in larger, ethnically diverse cohorts to improve the generalizability and robustness of the conclusions. The incorporation of tissue-specific mQTL data, particularly from buccal cells, would enhance the consistency of the analyses and better reflect tissue-specific contexts. Methodologically, the use of alternative MR approaches, such as MR-Egger, MR-PRESSO, and the weighted median method, may help address potential violations of the core assumptions and increase the reliability of causal inference [27]. Where possible, bidirectional MR analyses should also be conducted to assess the potential for reverse causation. In addition, functional follow-up studies, such as epigenetic editing or chromatin interaction assays, are warranted to validate the regulatory roles of key DNAm loci identified in this study. In addition to mental health outcomes, similar study framework can be applied to study DNAm as a potential mechanism linking PAE with physiological health outcomes. Furthermore, given the heterogeneity in epigenetic effects across different drinking patterns and gestational time windows, future studies should adopt more refined exposure characterizations to better understand the timing-specific and dose-dependent impacts of prenatal alcohol exposure on mental health risk [56]. Lastly, given the difference of CpG sites and discovery rates between the Lussier study and Sharp study, future study can compare impacts of PAE on health outcomes with different DNAm measuring time points, such as at birth, during childhood, and in adulthood.

## CONCLUSIONS

This study provided evidence that DNAm may be a mechanism that links PAE to adverse mental health outcomes. SCZ, BD, and depression had the most associated DNAm loci. The direction and strength of these associations varied by the timing and pattern of PAE exposure. The nominal DNAm loci may involve in developmental and neural functions. Together, the results highlighted DNAm as a key mechanism in the etiology of PAE-related psychiatric risks, which may ultimately lead to novel intervention and therapeutic strategies.

## ACKNOWLEDGEMENTS

This work was supported by a grant from the NIAAA (R21AA030640). The content is solely the responsibility of the authors and does not necessarily represent the official views of the National Institutes of Health. AAL is supported by an MQ Fellows Award (MQF22\9). The Substance Use Disorders Working Group of the Psychiatric Genomics Consortium (PGC-SUD) is supported by funds from NIDA and NIMH to MH109532. We gratefully acknowledge our contributing studies and the participants in those studies without whom this effort would not be possible. The funders took no role in the design, execution, analysis or interpretation of the data or in the writing up of the findings.

## DATA AVAILABILITY

Summary statistics were obtained from publicly-available sources. Summary statistics for DNAm loci were downloaded from http://godmc.org.uk/. Summary statistics for ADHD, anxiety disorders, autism spectrum disorder, bipolar disorder, cannabis use disorder, opioid dependence, post-traumatic stress disorder (PTSD), and schizophrenia can be downloaded from https://pgc.unc.edu/. Summary statistics for depression can be downloaded from https://ipsych.dk/en/research/downloads. Summary statistics from suicide attempt can be downloaded from https://tinyurl.com/ISGC2021.

