## Supplemental figures for "Prenatal Alcohol Exposure and Mental Health Outcomes: A Two-Sample Mendelian Randomization Study of DNA Methylation Signatures"

**Supplementary Figures**

**Figure S1.** Annotated forest plots for associations across DNAm loci and mental health outcomes (Lussier study).
**Figure S2.** Forest plot for associations between DNAm loci and 3 AUD traits (Lussier study)
**Figure S3.** Annotated forest plots for associations across DNAm loci and mental health outcomes (Sharp study).

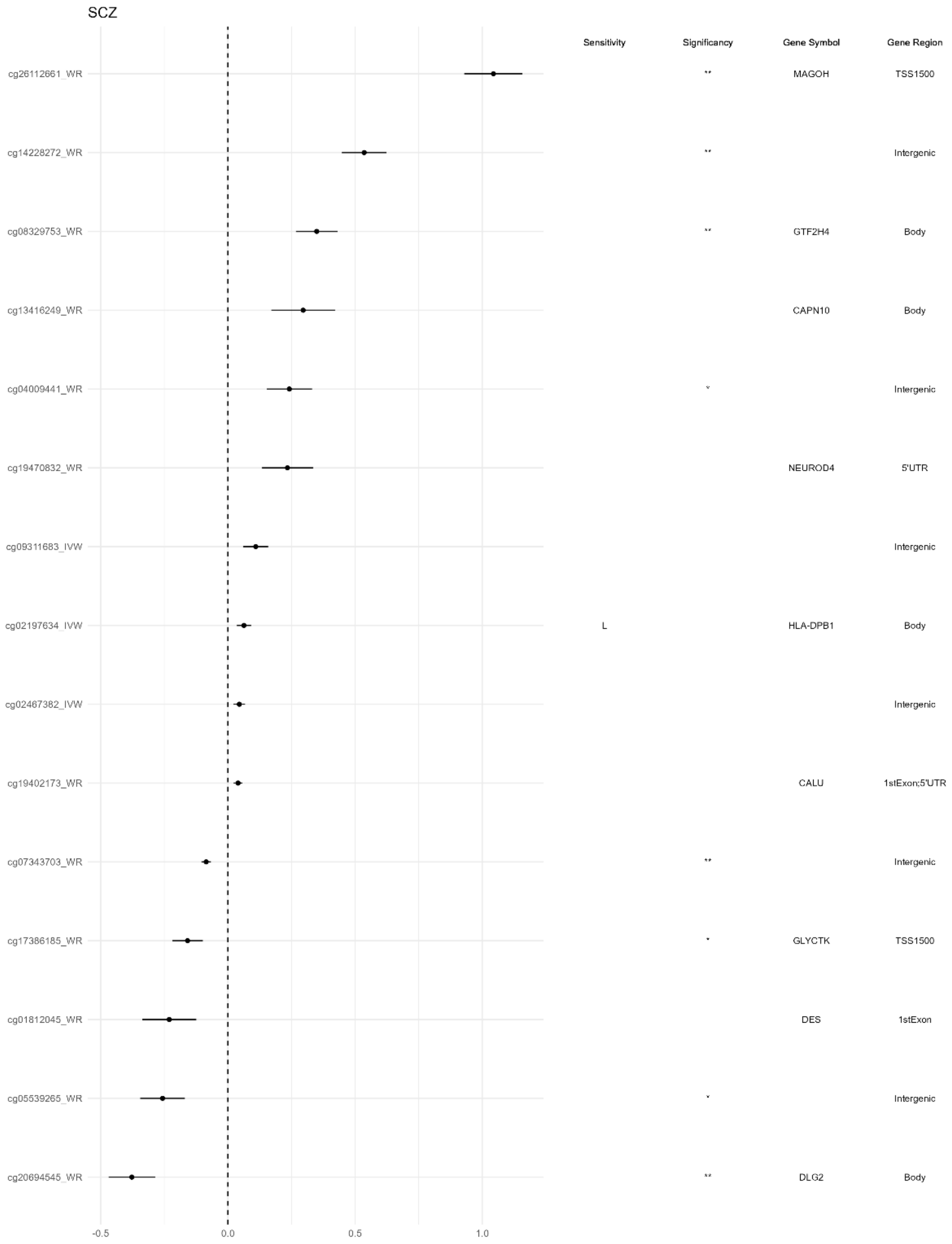
SCZ
Sensitivity
Significancy
Gene Symbol
Gene Region
cg26112661_WR
**
MAGOH
TSS1500
cg14228272_WR
**
Intergenic
cg08329753_WR
**
GTF2H4
Body
cg13416249_WR
CAPN10
Body
cg04009441_WR
*
Intergenic
cg19470832_WR
NEUROD4
5'UTR
cg09311683_IVW
Intergenic
cg02197634_IVW
L
HLA-DPB1
Body
cg02467382_IVW
Intergenic
cg19402173_WR
CALU
1stExon;5'UTR
cg07343703_WR
**
Intergenic
cg17386185_WR
*
GLYCTK
TSS1500
cg01812045_WR
DES
1stExon
cg05539265_WR
*
Intergenic
cg20694545_WR
**
DLG2
Body
-0.5
0.0
0.5
1.0

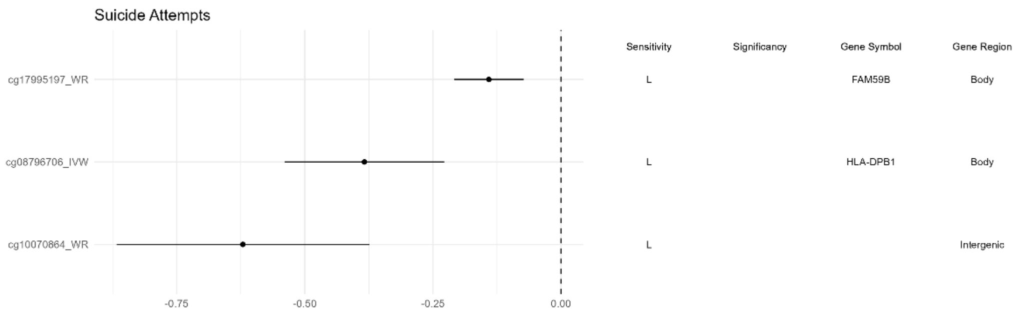
Suicide Attempts
Sensitivity
Significancy
Gene Symbol
Gene Region
cg17995197_WR
L
FAM59B
Body
cg08796706_IVW
L
HLA-DPB1
Body
cg10070864_WR
L
Intergenic
-0.75
-0.50
-0.25
0.00

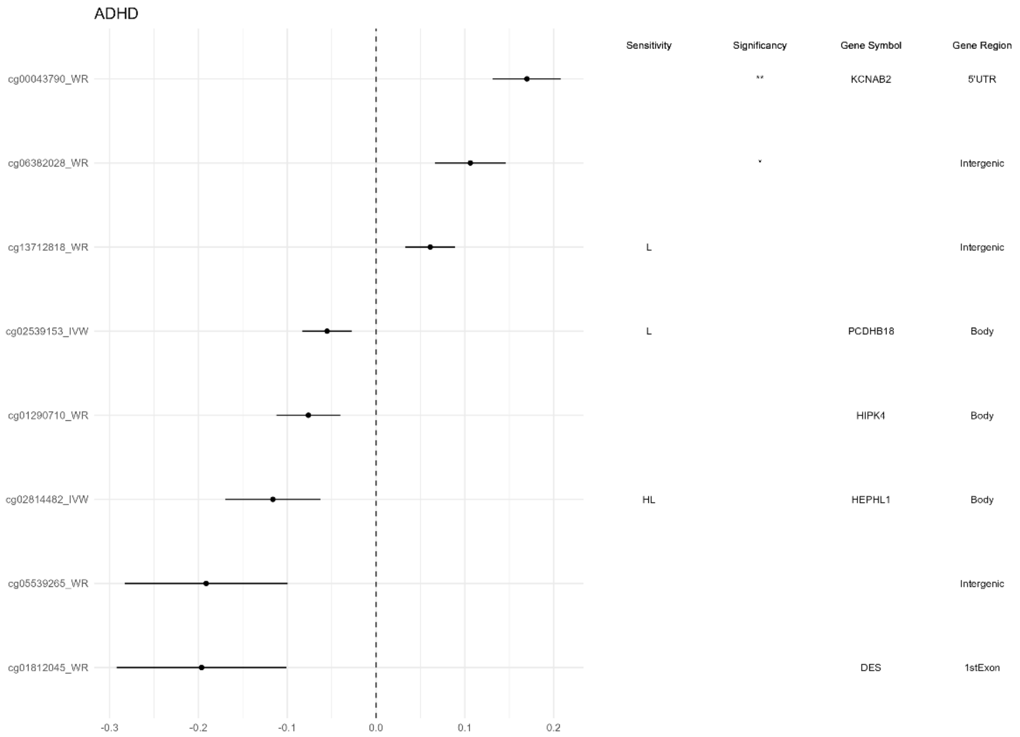
ADHD
Sensitivity
Significancy
Gene Symbol
Gene Region
cg00043790_WR
**
KCNAB2
5'UTR
cg06382028_WR
*
Intergenic
cg13712818_WR
L
Intergenic
cg02539153_IVW
L
PCDHB18
Body
cg01290710_WR
HIPK4
Body
cg02814482_IVW
HL
HEPHL1
Body
cg05539265_WR
Intergenic
cg01812045_WR
DES
1stExon
-0.3
-0.2
-0.1
0.0
0.1
0.2

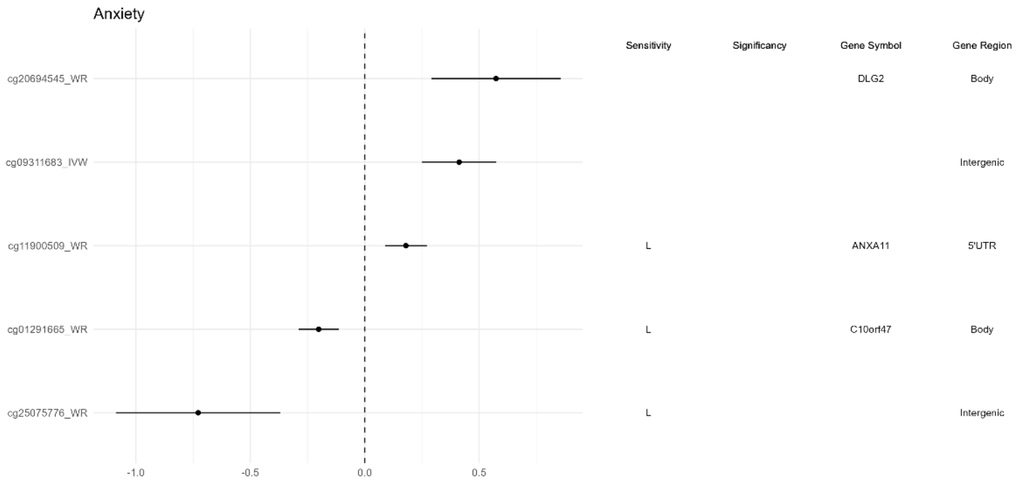
Anxiety
Sensitivity
Significancy
Gene Symbol
Gene Region
cg20694545_WR
DLG2
Body
cg09311683_IVW
Intergenic
cg11900509_WR
L
ANXA11
5'UTR
cg01291665_WR
L
C10orf47
Body
cg25075776_WR
L
Intergenic
-1.0
-0.5
0.0
0.5

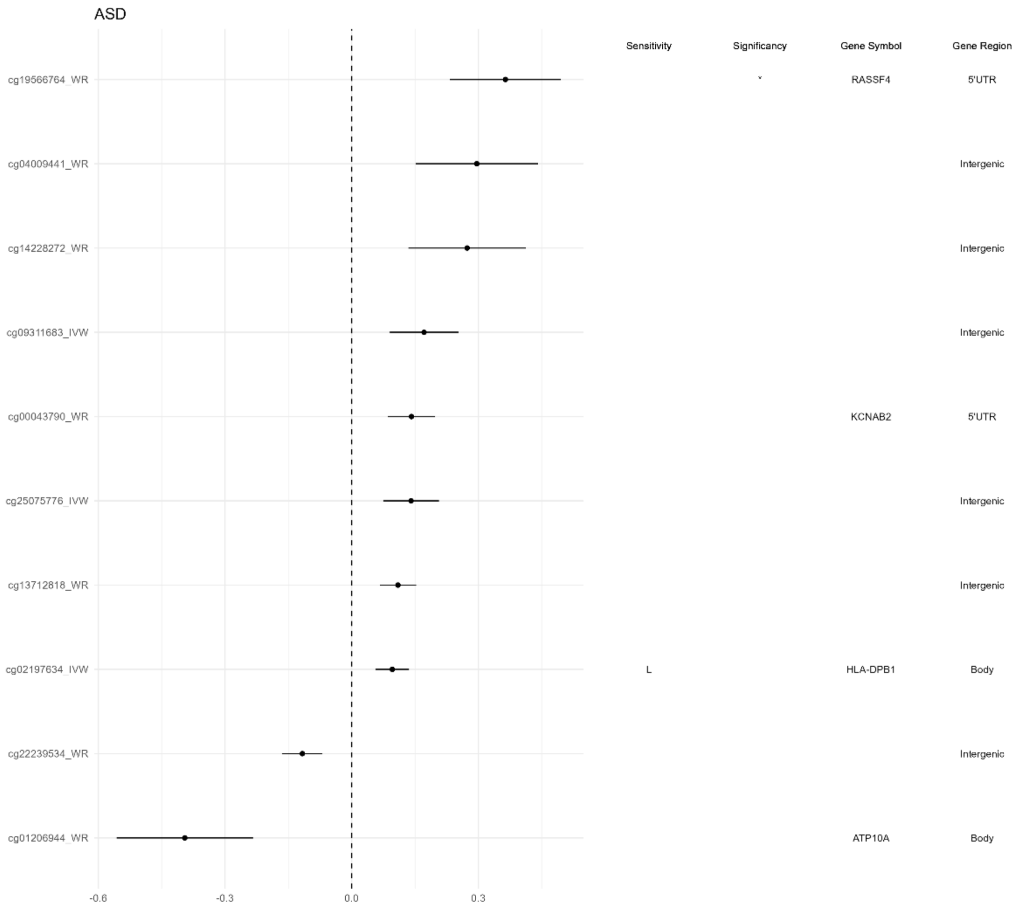
ASD
Sensitivity
Significancy
Gene Symbol
Gene Region
cg19566764_WR
*
RASSF4
5'UTR
cg04009441_WR
Intergenic
cg14228272_WR
Intergenic
cg09311683_IVW
Intergenic
cg00043790_WR
KCNAB2
5'UTR
cg25075776_IVW
Intergenic
cg13712818_WR
Intergenic
cg02197634_IVW
L
HLA-DPB1
Body
cg22239534_WR
Intergenic
cg01206944_WR
ATP10A
Body
-0.6
-0.3
0.0
0.3

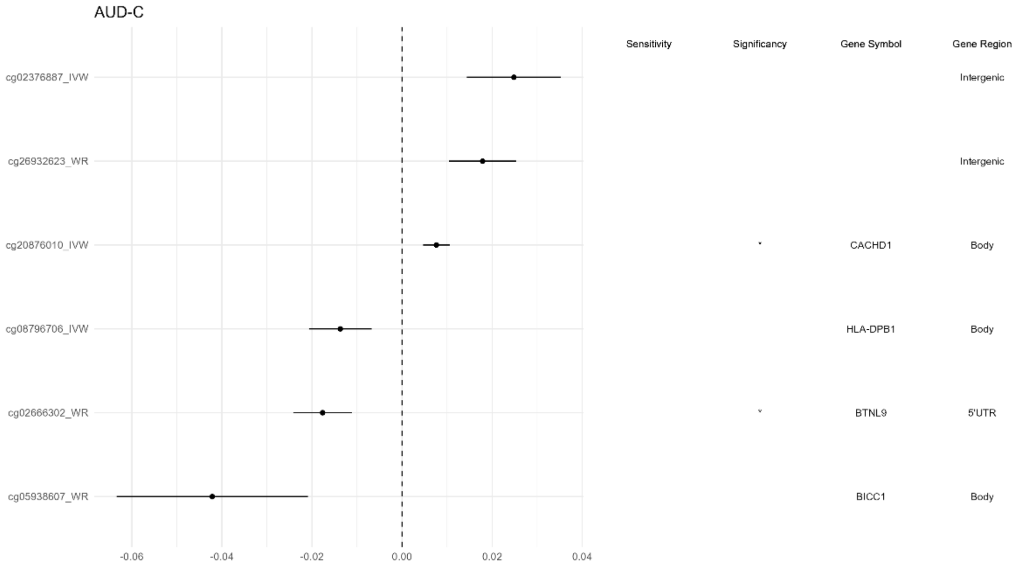
AUD-C
Sensitivity
Significancy
Gene Symbol
Gene Region
cg02376887_IVW
Intergenic
cg26932623_WR
Intergenic
cg20876010_IVW
*
CACHD1
Body
cg08796706_IVW
HLA-DPB1
Body
cg02666302_WR
*
BTNL9
5'UTR
cg05938607_WR
BICC1
Body
-0.06
-0.04
-0.02
0.00
0.02
0.04

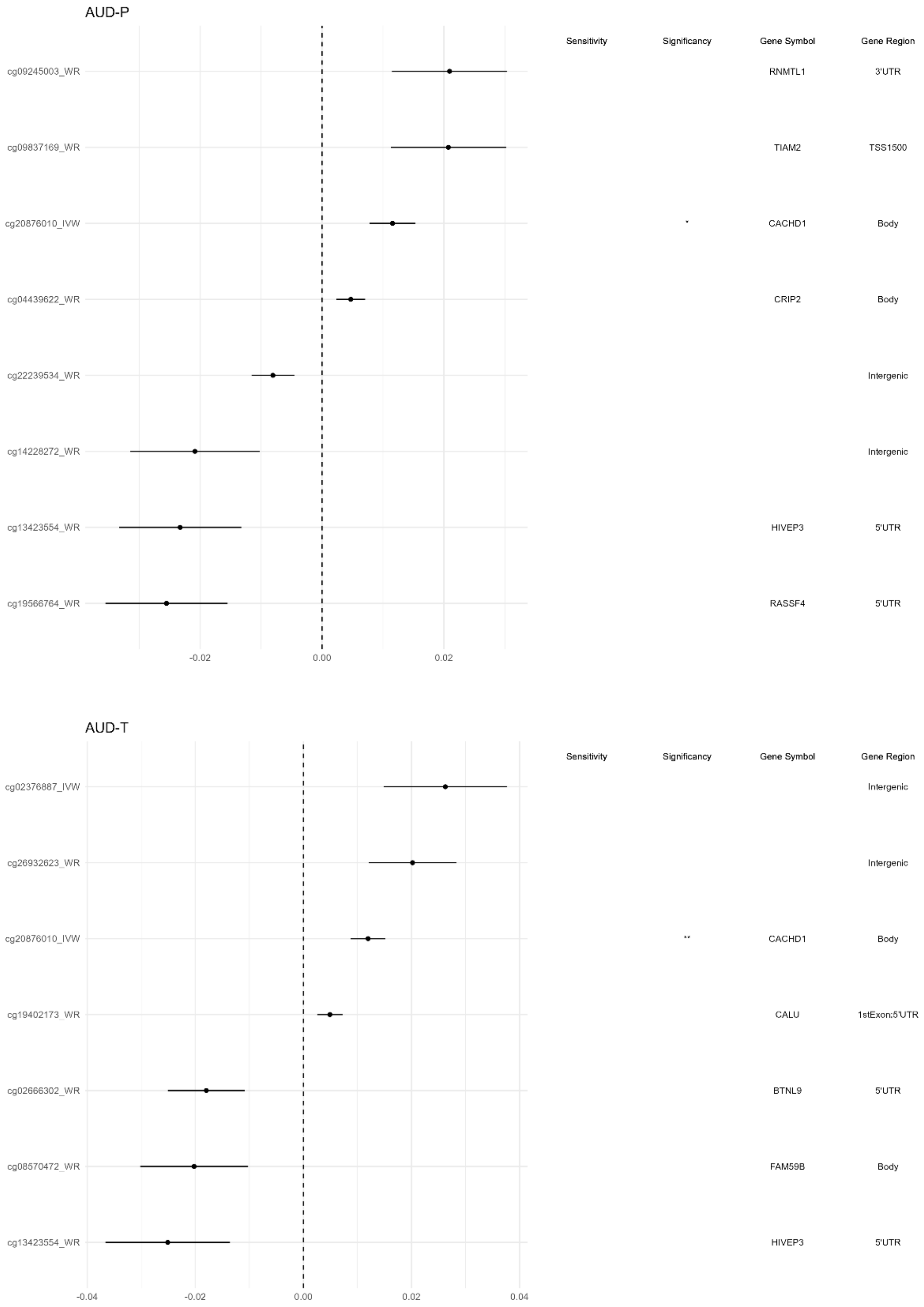
AUD-P
Sensitivity
Significancy
Gene Symbol
Gene Region
cg09245003_WR
RNMTL1
3'UTR
cg09837169_WR
TIAM2
TSS1500
cg20876010_IVW
*
CACHD1
Body
cg04439622_WR
CRIP2
Body
cg22239534_WR
Intergenic
cg14228272_WR
Intergenic
cg13423554_WR
HIVEP3
5'UTR
cg19566764_WR
RASSF4
5'UTR
-0.02
0.00
0.02
AUD-T
Sensitivity
Significancy
Gene Symbol
Gene Region
cg02376887_IVW
Intergenic
cg26932623_WR
Intergenic
cg20876010_IVW
**
CACHD1
Body
cg19402173_WR
CALU
1stExon;5'UTR
cg02666302_WR
BTNL9
5'UTR
cg08570472_WR
FAM59B
Body
cg13423554_WR
HIVEP3
5'UTR
-0.04
-0.02
0.00
0.02
0.04

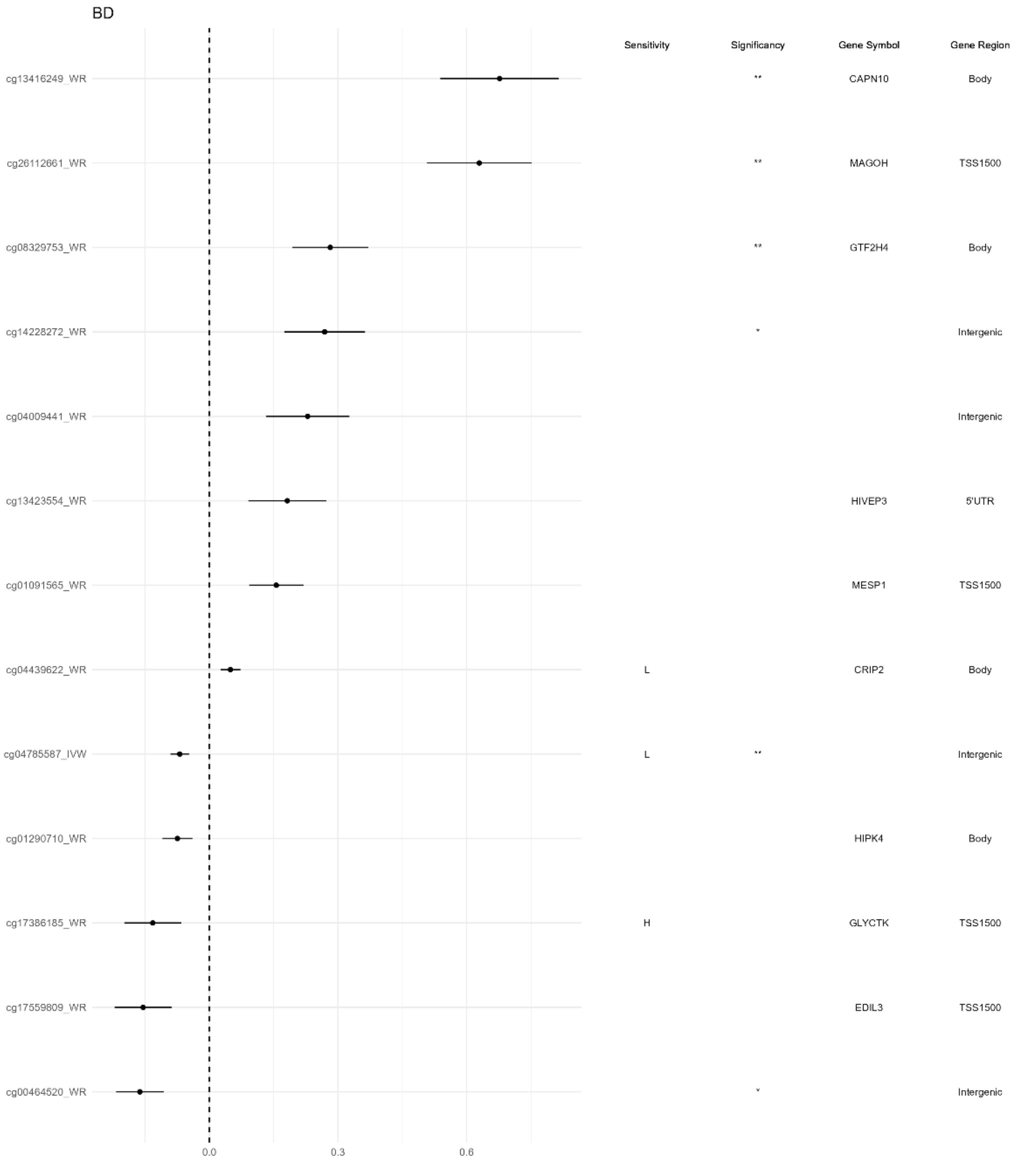
BD
Sensitivity
Significancy
Gene Symbol
Gene Region
cg13416249_WR
**
CAPN10
Body
cg26112661_WR
**
MAGOH
TSS1500
cg08329753_WR
**
GTF2H4
Body
cg14228272_WR
*
Intergenic
cg04009441_WR
Intergenic
cg13423554_WR
HIVEP3
5'UTR
cg01091565_WR
MESP1
TSS1500
cg04439622_WR
L
CRIP2
Body
cg04785587_IVW
L
**
Intergenic
cg01290710_WR
HIPK4
Body
cg17386185_WR
H
GLYCTK
TSS1500
cg17559809_WR
EDIL3
TSS1500
cg00464520_WR
*
Intergenic
0.0
0.3
0.6

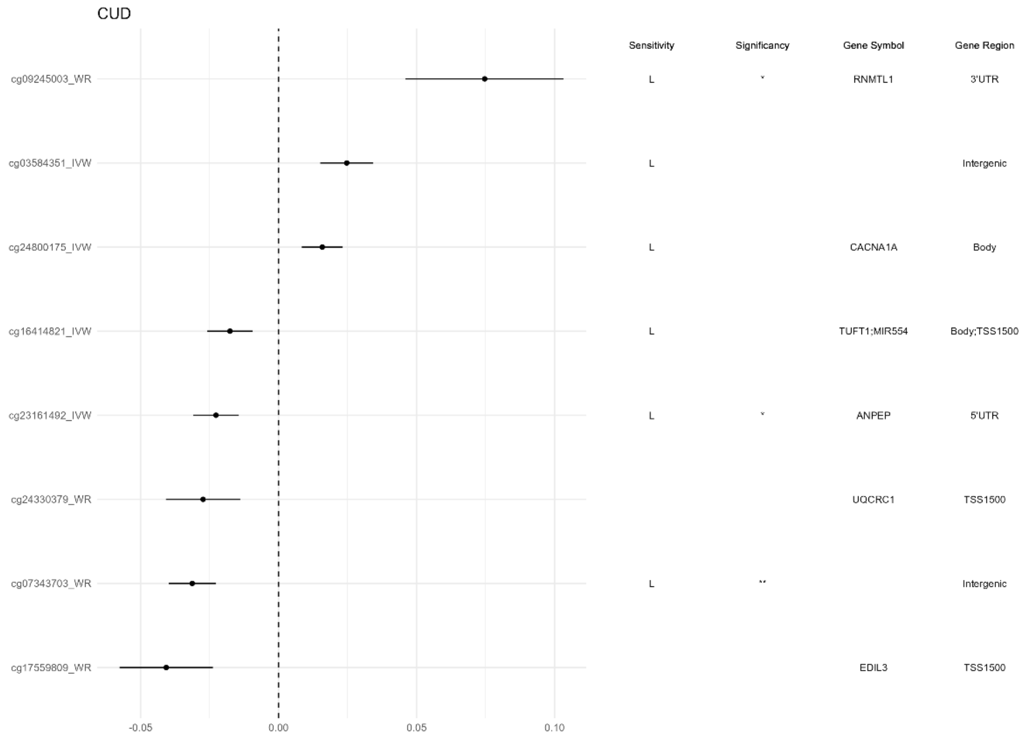
CUD
Sensitivity
Significancy
Gene Symbol
Gene Region
cg09245003_WR
L
*
RNMTL1
3'UTR
cg03584351_IVW
L
Intergenic
cg24800175_IVW
L
CACNA1A
Body
cg16414821_IVW
L
TUFT1;MIR554
Body;TSS1500
cg23161492_IVW
L
*
ANPEP
5'UTR
cg24330379_WR
UQCRC1
TSS1500
cg07343703_WR
L
**
Intergenic
cg17559809_WR
EDIL3
TSS1500
-0.05
0.00
0.05
0.10

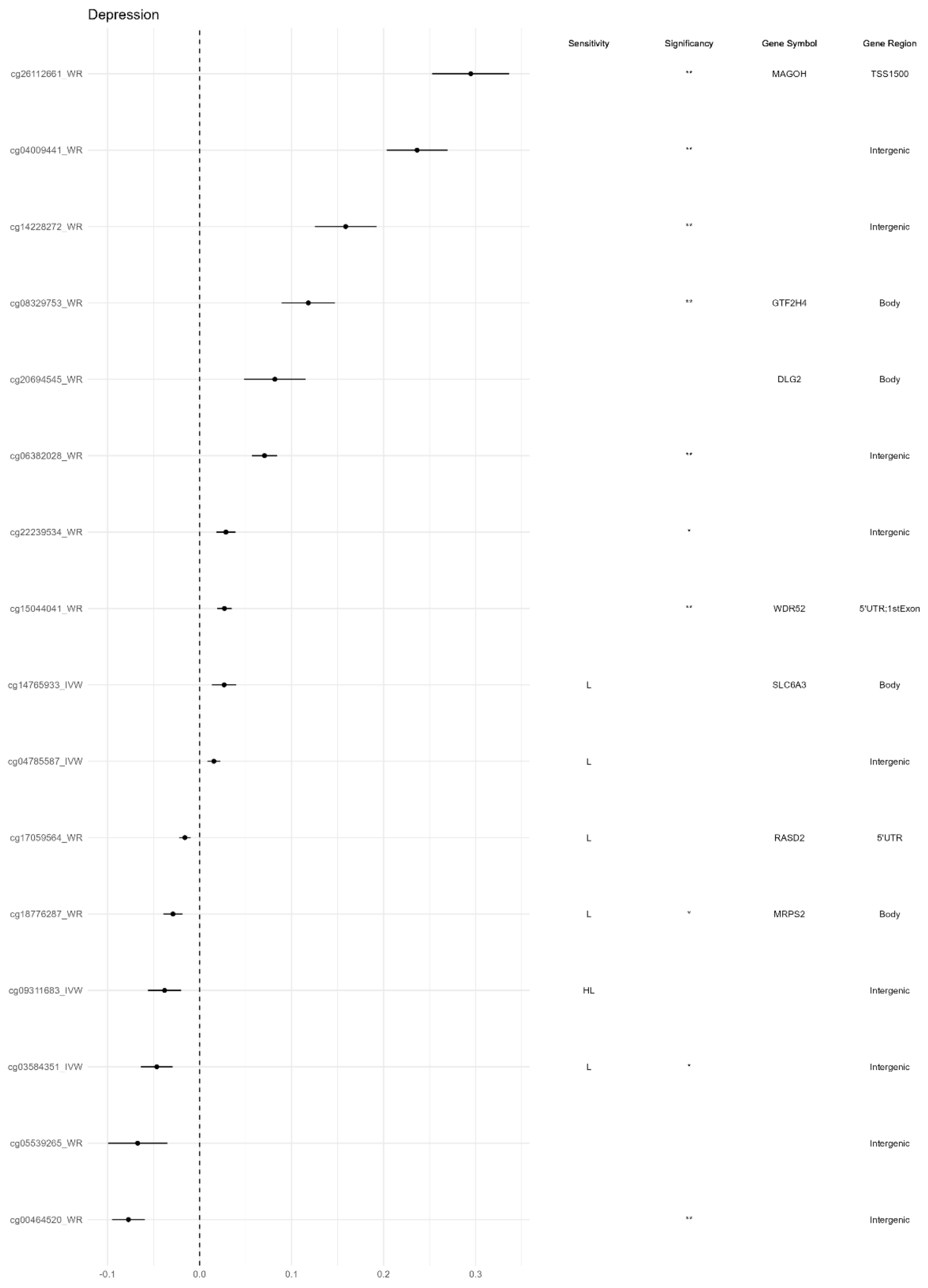
Depression
Sensitivity
Significancy
Gene Symbol
Gene Region
cg26112661_WR
**
MAGOH
TSS1500
cg04009441_WR
**
Intergenic
cg14228272_WR
**
Intergenic
cg08329753_WR
**
GTF2H4
Body
cg20694545_WR
DLG2
Body
cg06382028_WR
**
Intergenic
cg22239534_WR
*
Intergenic
cg15044041_WR
**
WDR52
5'UTR;1stExon
cg14765933_IVW
L
SLC6A3
Body
cg04785587_IVW
L
Intergenic
cg17059564_WR
L
RASD2
5'UTR
cg18776287_WR
L
*
MRPS2
Body
cg09311683_IVW
HL
Intergenic
cg03584351_IVW
L
*
Intergenic
cg05539265_WR
Intergenic
cg00464520_WR
**
Intergenic
-0.1
0.0
0.1
0.2
0.3

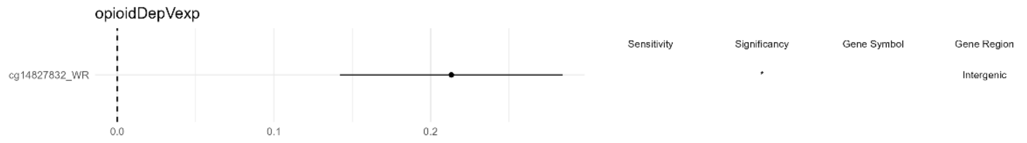
opioidDepVexp
Sensitivity
Significancy
Gene Symbol
Gene Region
cg14827832_WR
*
Intergenic
0.0
0.1
0.2

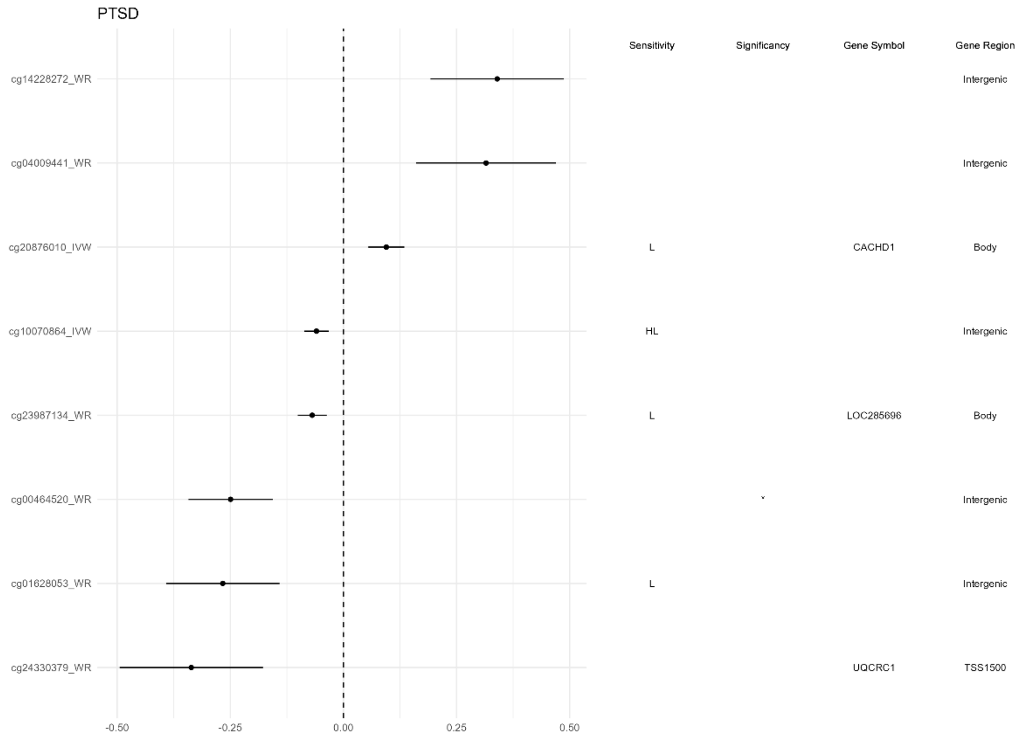


**Figure S1.** Annotated forest plots for associations across DNAm loci and mental health outcomes (Lussier study). Each sub-plot corresponds to one mental health outcome. Right to each forest plot, there are 4 annotation columns: Sensitivity (empty means passing all sensitivity tests; "H" means failing the heterogeneity test; "P" means failing the pleiotropy test; "L" means failing the leave-one-out test), Significancy (*pval<.01; **FDR<.05), mapped gene symbols, and mapped gene regions.

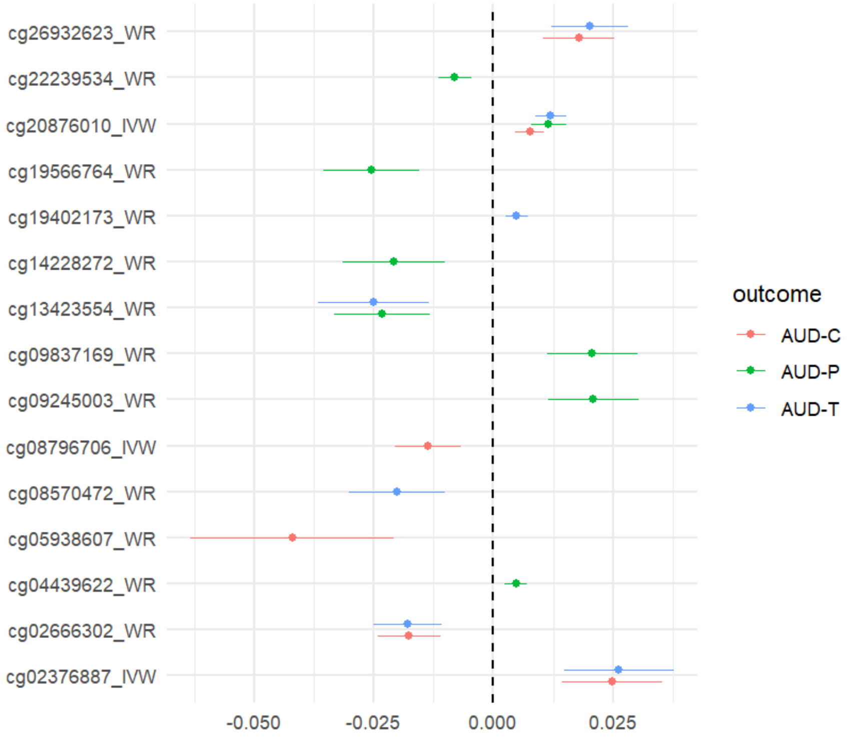


**Figure S2.** Forest plot for associations between DNAm loci and 3 AUD traits (Lussier study).

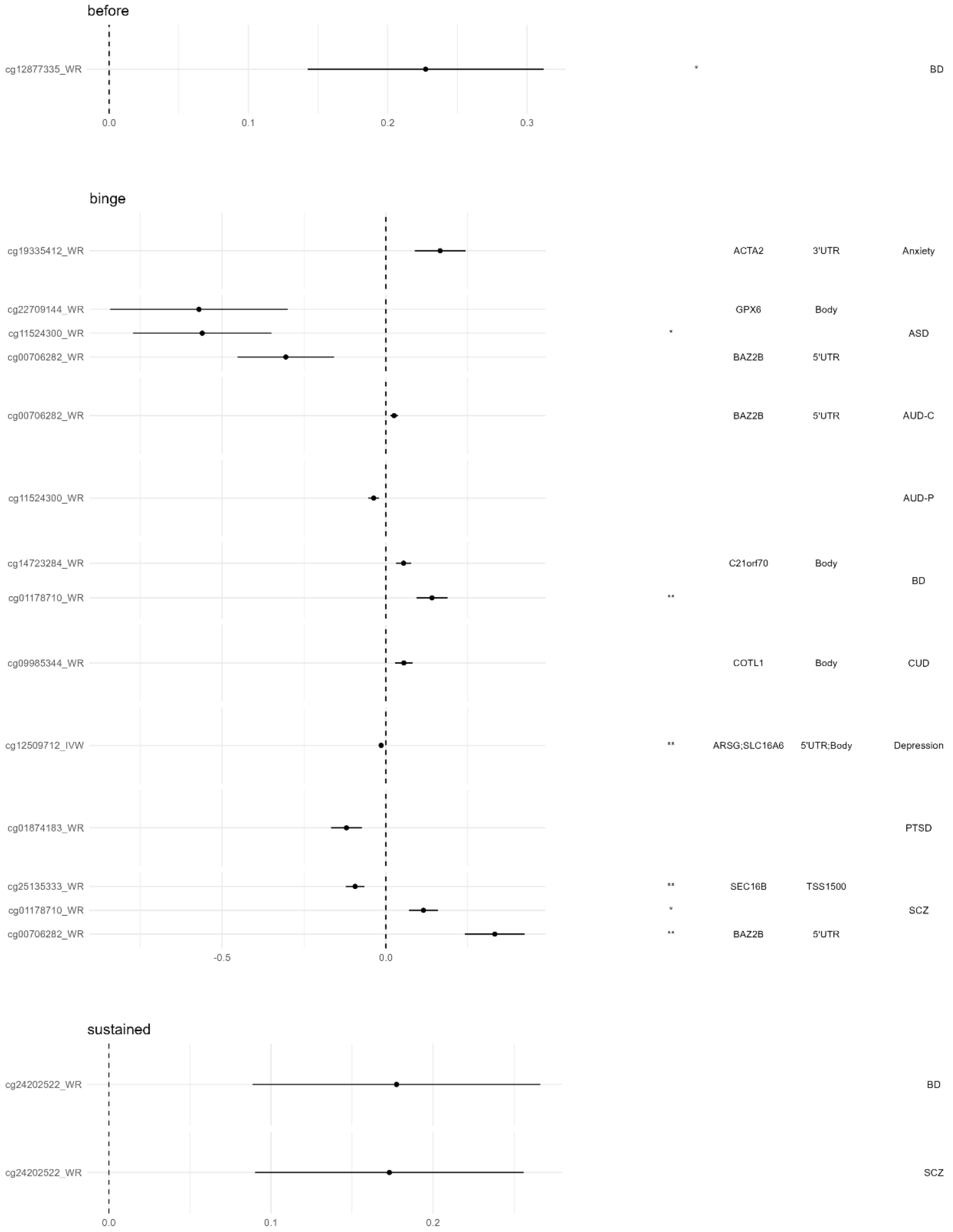

before
cg12877335_WR
*
BD
0.0
0.1
0.2
0.3
binge
cg19335412_WR
ACTA2
3'UTR
Anxiety
cg22709144_WR
GPX6
Body
cg11524300_WR
*
ASD
cg00706282_WR
BAZ2B
5'UTR
cg00706282_WR
BAZ2B
5'UTR
AUD-C
cg11524300_WR
AUD-P
cg14723284_WR
C21orf70
Body
BD
cg01178710_WR
**
cg09985344_WR
COTL1
Body
CUD
cg12509712_IVW
**
ARSG;SLC16A6
5'UTR;Body
Depression
cg01874183_WR
PTSD
cg25135333_WR
**
SEC16B
TSS1500
cg01178710_WR
*
SCZ
cg00706282_WR
**
BAZ2B
5'UTR
-0.5
0.0
sustained
cg24202522_WR
BD
cg24202522_WR
SCZ
0.0
0.1
0.2

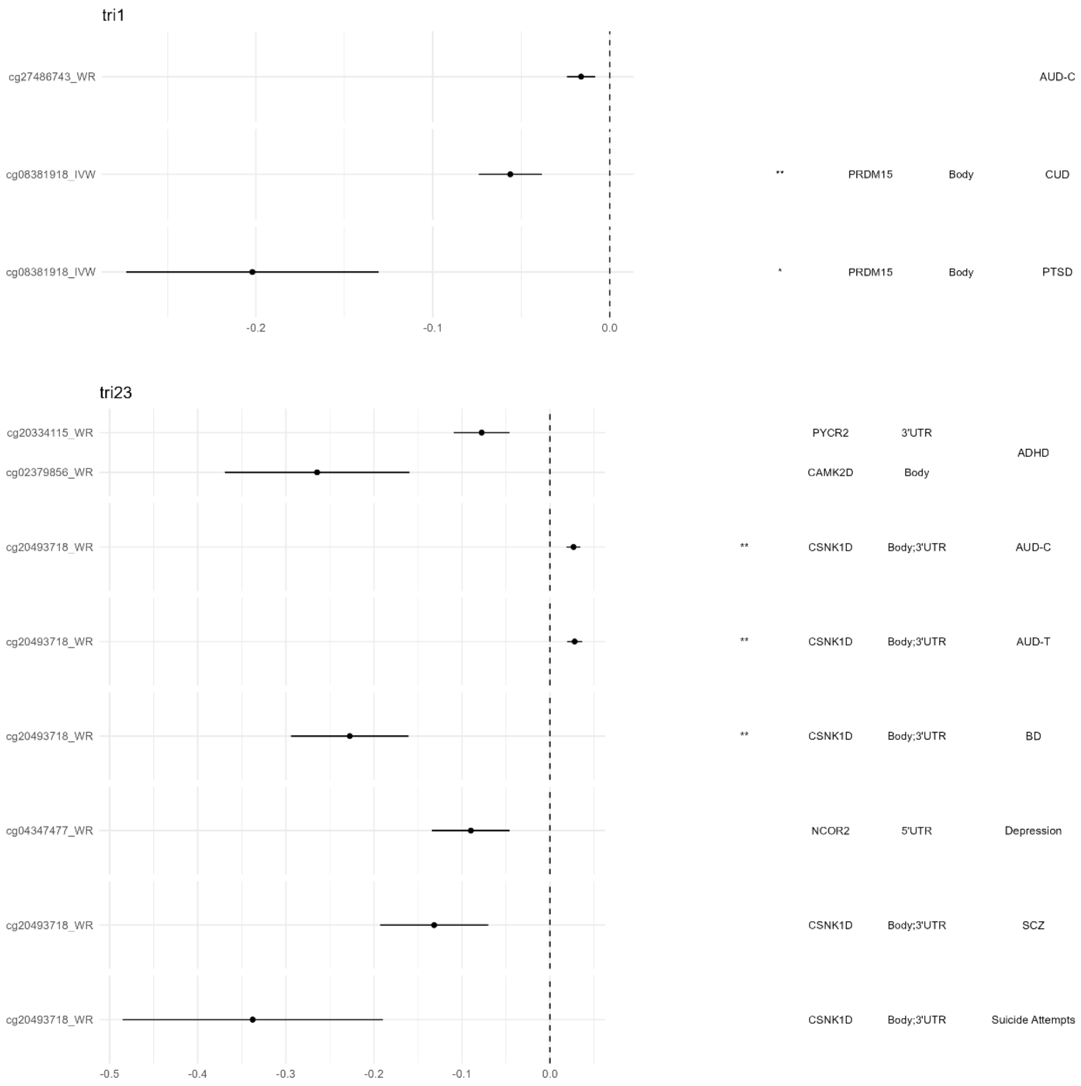


**Figure S3.** Annotated forest plots for associations across DNAm loci and mental health outcomes (Sharp study). Each sub-plot corresponds to one drinking pattern. Right to each forest plot, there are 4 annotation columns: Sensitivity (empty means passing all sensitivity tests), Significance (*pval<.01; **FDR<.05), mapped gene symbols, and mapped gene regions. The right most column denotes the mental health outcomes for each sub-layer in each sub-plot.
